# Influence of Prior SARS-CoV-2 Infection on COVID-19 Severity: Evidence from the National COVID Cohort Collaborative

**DOI:** 10.1101/2023.08.03.23293612

**Authors:** Nathaniel Hendrix, Hythem Sidky, David K. Sahner, The N3C Consortium

## Abstract

**Background:** A large share of SARS-CoV-2 infections now occur among previously infected individuals. In this study, we sought to determine whether prior infection modifies disease severity relative to no prior infection.

**Methods:** We used data from first and second COVID-19 episodes in the National COVID Cohort Collaborative, a nationwide collection of de-identified electronic health records. We used nested logistic regressions of monthly cohorts weighted on the inverse probability of prior infection to assess risk of hospitalization, death, and increased severity in the first versus second infection cohorts.

**Results:** We included a total of 2,058,274 individuals in the analysis, 147,592 of whom had two recorded infections. The impact of prior infection differed meaningfully between months. Prior infection was largely protective prior to March 2022, with odds ratios (ORs) as low as 0.66 (95% confidence interval: 0.51 to 0.86) in November 2021 for hospitalization. and as low as 0.23 (0.06 to 0.86) in June 2021 for death. However, prior infection was associated with an increased risk of hospitalization and death, mostly after March 2022 when the ORs were as high as 1.87 (1.26 to 2.80) and 2.99 (1.65 to 5.41) in April 2022, respectively. The overall OR for more severe disease was 1.06 (1.03 to 1.10) among previously infected individuals.

**Conclusion:** In the pandemic’s first two years, previously infected patients generally had less severe disease than people without prior infection. During the Omicron era, however, previously infected patients had the same or worse severity of disease as patients without prior infection.

## Introduction

The SARS-CoV-2 pandemic’s ongoing shift towards endemic status has necessarily been accompanied by substantial adjustments to public health policy. Recommendations for vaccine boosters, public investment in novel treatments for COVID-19, and reimbursement for telemedicine have already changed or begun to be debated.(1) As of late 2022, a substantial majority of Americans had experienced at least one episode of COVID-19.(2) As more individuals become reinfected, an understanding of how prior infection changes disease severity is vital to making policy decisions informed by reasonable risk-benefit calculations.

One of the hallmarks of an infectious disease’s transition from epidemic to endemic status is a reduction in disease severity due, in part, to an increasing number of the population with some immune protection against the disease.(3) In many cases, this takes place through prior infections. It is therefore reasonable to hypothesize that prior infection with SARS-CoV-2 also confers some protective effect. Some evidence, though, has suggested that the protective effect of natural antibodies against SARS-CoV-2 may decay relatively quickly compared to antibodies against other viruses.(4) Published efforts to understand how repeat COVID-19 affects disease severity have been surprisingly rare, in part due to the notable challenges of studying repeat infection.(5) Among other challenges, these have included the emergence of several new variants since the start of the pandemic, the introduction of vaccines, and the likely increase in the rate of self-management for COVID-19.

In this study, we used the National COVID Cohort Collaborative (N3C) dataset to conduct an analysis of how prior infection modifies the risk of severe COVID-19 outcomes. The N3C collected deidentified data from 76 health systems covering 18.9 million individuals and 7.5 million COVID-19 cases to provide comprehensive information on a large, diverse swath of patients throughout the United States.(6) We used causal methods with these data to assess the relevance of reinfection to COVID-19 severity both at the population level and across subgroups defined by gender, race/ethnicity, and age.

## Methods

### Data Source

The National Institutes of Health (NIH) sponsored the creation of the N3C dataset, which is the largest collection of information on COVID-19 infections in the United States. The N3C uses electronic health record data volunteered from healthcare organizations on patients in their systems going as far back as January 1, 2018 and extending to March 31, 2023. The data are cleaned and then harmonized into a single data standard using the Observational Medical Outcomes Partnership (OMOP) ontology. The harmonized data are then hosted in a secure online portal for researchers. Episodes of COVID-19 were defined by the N3C COVID-19 Phenotype, v4.0.(7) This phenotype required either a positive lab test (PCR or antigen) or diagnosis code specific to COVID-19. For cases prior to May 1, 2020, at which point diagnostic testing was not widely available, the phenotype allowed for identification of COVID-19 via two weak positive diagnostic codes documented on the same day. These weak positive codes included exposure to virus and symptoms.

### Cohort Identification

We included only individuals who had at least one recorded COVID-19 infection in our analysis. All included patients were required to have at least one visit in the year prior to their first recorded infection. This lookback period increased our confidence in the availability of baseline medical history, including 32 comorbidities with probable associations to COVID-19 outcomes (Supplemental Material I).(8) Vaccination is known to be a major determinant of COVID-19 outcomes, yet a large share of vaccination took place in community settings that are not well-integrated into most EHRs. We included data from eight health systems who had been established to have vaccination rates that generally matched with their locale’s overall vaccination rate and, thus, likely had the best integration of vaccination data into their EHRs.(9)

We defined reinfection as the first infection to take place at least 60 days after the earliest recorded infection. To further ensure that participant privacy was protected, we only included months with a total of at least 20 reinfections.

### Risk of Hospitalization and Death

Our first analysis assessed two outcomes: hospitalization and death. We analyzed data on these two disease outcomes as if it were produced by a sequential series of nested trials, each starting within a given month.

Thus, within each month with at least 20 reinfections, we compared the risks of hospitalization and death of those with and without prior COVID-19. This approach allowed us to manage the challenges associated with the multitude of time-varying influences on severity throughout the pandemic.(10) Our outcomes of interest were the counterfactual distributions hospitalization and death among the cohort of those with a second infection at the time of infection had it instead been their first.

We analyzed the observational data using a two-stage approach. In the first stage, we used a logistic regression within each month to predict the probability that the individual had a prior recorded episode of COVID-19. The regression included the following independent variables: age, age squared, race/ethnicity, sex, health system data source, whether the patient’s postal code was recorded, count of vaccinations having taken place at least 2 weeks prior to the infection date (categorical from 0 to more than 4), and body mass index (categorical as <18.5, 18.5 - 24.9, 25 - 29.9, 30 - 34.9, 35 – 40, >40, or missing). We also included in the regression a binary indicator for each of the comorbidities listed above. We converted the predicted probability of prior exposure into stabilized inverse probability of treatment weights. The balance of potentially outcome-relevant covariates was confirmed by plots of standardized mean differences using a threshold of 0.1 (Supplemental Material II).

In the second stage, we used two logistic regressions to identify the effect of prior infection on the risks of hospitalization and death. We estimated the monthly effect of exposure by including an interaction term between the binary variable for prior infection and a categorical variable for the month. In both regressions, each patient was weighted by the inverse probability of treatment weights calculated in the first stage of the analysis, thereby also accounting for covariates that affect disease severity. We also pooled the monthly results into an estimate of pandemic-wide risk of severe disease by repeating the analysis without terms for the month of infection.

Our analysis hinged on several important assumptions. We tested the relevance of these assumptions to our findings on the risk of hospitalization and death through two sensitivity analyses. Namely, we first evaluated the impact of prior infection on a cohort without the one-year lookback period required in the main analysis. In another analysis, we changed the definition of a second infection from an infection taking place at least 60 days after initial infection to one taking place at least 90 days later. In both cases, we repeated the above analyses for the risk of hospitalization and death but with modified criteria for cohort and reinfection criteria.

### Risk of More Severe Disease

Our analysis of hospitalization and death may disguise finer-grained distinctions of severity, such as the distinction between hospitalized patients who were and were not ventilated. Therefore, we also analyzed outcomes using the five-level classification of COVID-19 severity defined by N3C. The classifications were:

- Mild: Seen in an outpatient, non-emergent setting without subsequent hospitalization or death
- Mild in emergency department: Seen in emergency department without subsequent hospitalization or death
- Moderate: Hospitalized without extracorporeal membrane oxygenation (ECMO), invasive mechanical ventilation (IMV), or death
- Severe: Hospitalized with ECMO and/or IMV, but without death
- Death: Discharge status to “death” or “hospice” after a hospitalization with COVID-19

Mortality data in the N3C Enclave was enriched through privacy-preserving record linkage to ancillary sources of death data, including supplemental entries (SSA), private obituary entries, and obit.com entries.

For this analysis, we repeated the calculation of monthly weights in the binary analysis but modified the regression strategy. Specifically, we used an ordinal logistic regression to identify the effect of prior infection on disease severity. We again estimated both monthly and pandemic-wide effects.

In addition to the finer distinctions of severity that this analysis allowed, it also allowed us to perform subgroup analyses, which were infeasible in the binary analyses due to small numbers of events in the previously infected cohorts of some subgroups. We examined the effect of prior infection for the following subgroups:

- Black / African American, non-Hispanic
- Hispanic of any race
- White, non-Hispanic
- Female
- Male
- Age less than 18 years
- Age 18-44 years
- Age 45-59 years
- Age 60-69 years
- Age 70 or more years

We recalculated the inverse probability weight in each subgroup analysis because of the differing reference groups in the subgroup versus population analyses.

## Results

### Cohort

From an initial cohort of 7,446,481 combined first and second infections, we excluded observations due to lack of visits during the look-back period, origination from a health system without vaccination rates comparable to locally reported rates (i.e., presumably invalid vaccine data related to incomplete capture in the EHR), or infection in a month with fewer than 20 reinfections. We arrived at a final cohort of 2,058,274 first infections and 147,592 second infections (Figure 1). January 2022 contained the highest number of both first and second infections, with 351,754 and 40,905, respectively (Supplemental Material III).

**Figure 1:**
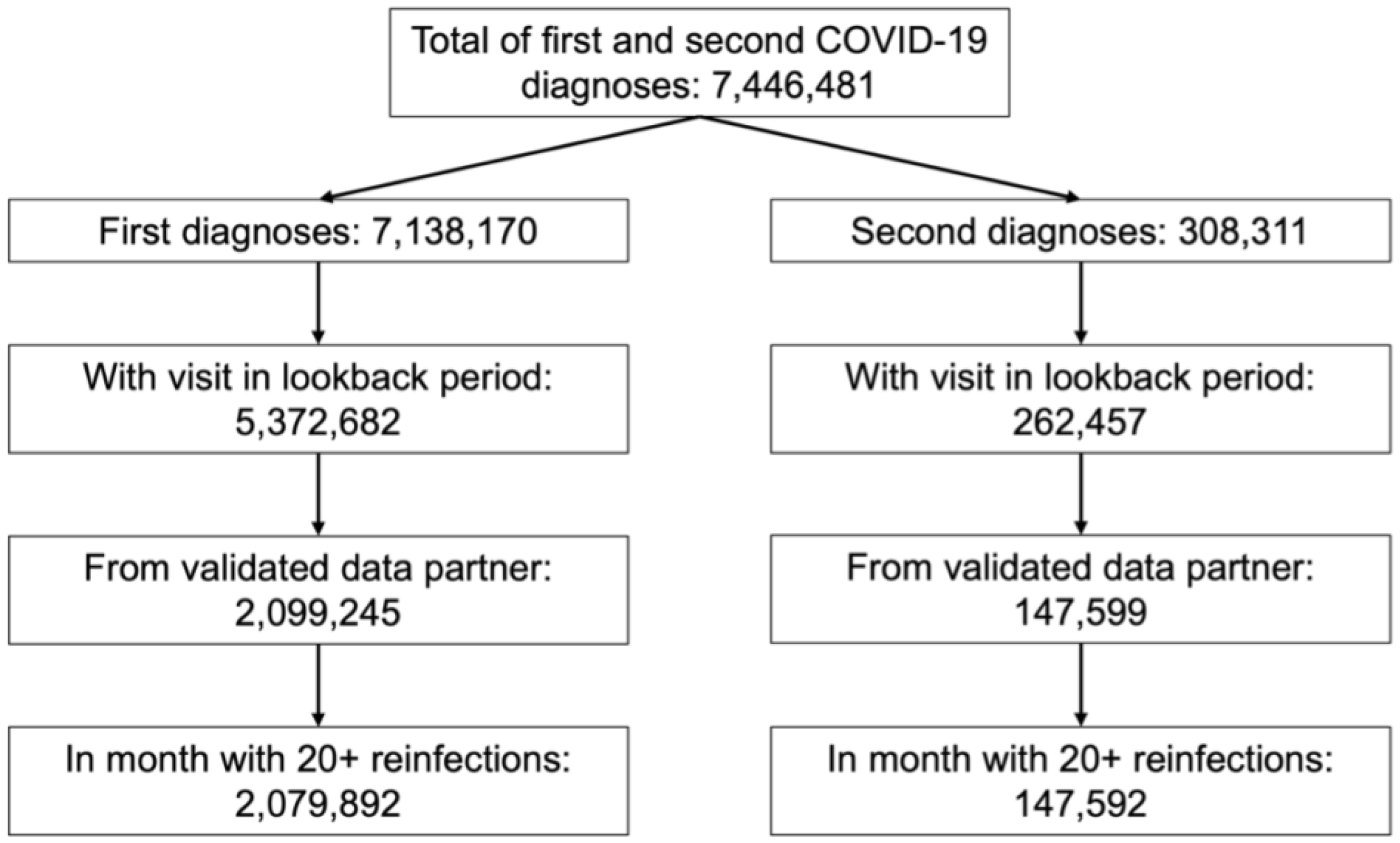
Data flow for the cohort included in the analysis.

Individuals with a recorded second infection were relatively similar to the cohort of individuals during their first infections (Table 1). The cohort with a recorded second infection was more likely to be female and somewhat more likely to be vaccinated, but overall had similar rates of comorbidities (Supplemental Material IV).

**Table 1:** Demographics at the time of first and second infection.

|  | First infections (n = 2,079,892) | Second infections (n = 147,592) |
| --- | --- | --- |
| Age at infection, mean (SD) | 42.1 (22.1) | 41.6 (20.2) |
| Sex: Female, n (%) | 1,195,238 (57.5%) | 95,961 (65.0%) |
| Sex: Male, n (%) | 882,972 (42.5%) | 51,825 (35.0%) |
| Race / ethnicity: Black, non-Hispanic, n (%) | 154,800 (7.4%) | 11,626 (7.9%) |
| Race / ethnicity: Hispanic, any race, n (%) | 103,374 (5.1%) | 4,493 (3.1%) |
| Race / ethnicity: White, non-Hispanic, n (%) | 1,502,224 (72.2%) | 109,979 (74.5%) |
| Race / ethnicity: Other, non-Hispanic, n (%) | 67,483 (3.2%) | 2,720 (1.8%) |

### Longitudinal Trends among Previously Reinfected Patients

At least two outcome-relevant characteristics of the cohort of previously infected individuals changed dramatically over the course of the pandemic (Figure 2). During 2020, the average time between first and second infection among this cohort was under 5 months, and vaccination against SARS-CoV-2 was minimal. During the first three months of 2023, however, first and second infections were 16 months apart on average, and approximately two-thirds of the reinfected cohort had at least one vaccination.

**Figure 2:**
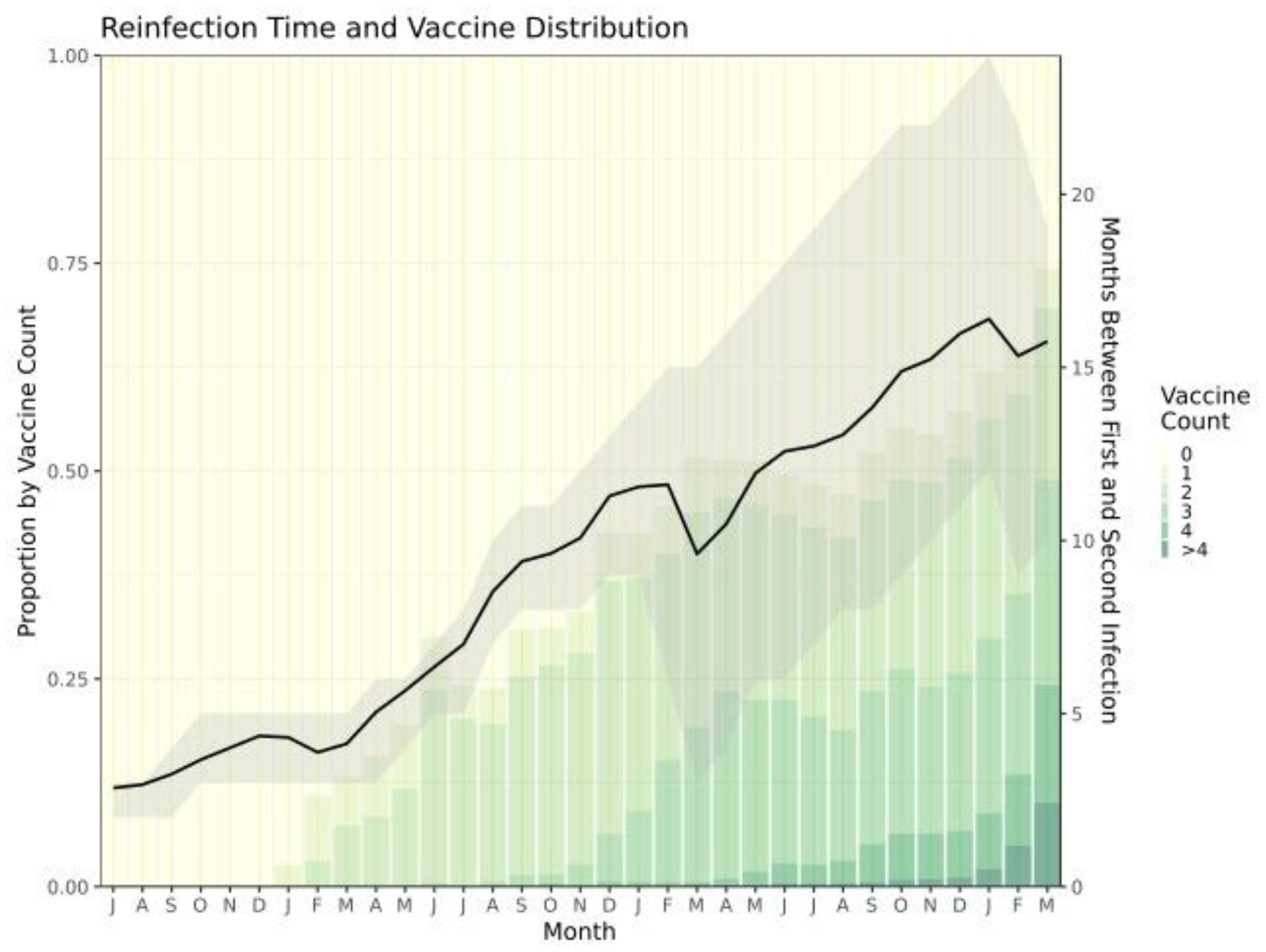
Longitudinal trends among the previously infected cohort in number of vaccines received and time between first and second infection. The shaded area represents the interquartile range of time between first and second infection.

### Reinfection Outcomes

Among patients with a recorded reinfection, 94% experienced mild disease during both infections. More severe outcomes were much rarer. Approximately 0.5% of reinfections resulted in death. Among individuals who had a severe initial infection, 5.9% died as a result of their second infection compared to much lower rates for individuals who had a less severe initial infection.

### Risks of Hospitalization and Death

Our binomial logistic regressions showed that prior infection’s effect on the risk of hospitalization and death relative to no prior infection was highly heterogeneous across the pandemic (Figure 3). There were four months, all prior to March 2022, in which prior infection provided a protective effect against hospitalization. There were nine months, largely in the period starting March 2022, in which prior infection was associated with a greater risk of hospitalization. Odds ratios (ORs) for hospitalization ranged from 0.66 (95% confidence interval [CI]: 0.51 to 0.86) in November 2021 to 1.87 (95% CI: 1.26 to 2.80) in April 2022. The pandemic-wide OR was 1.01 (95% CI: 0.96 to 1.07).

**Figure 3:**
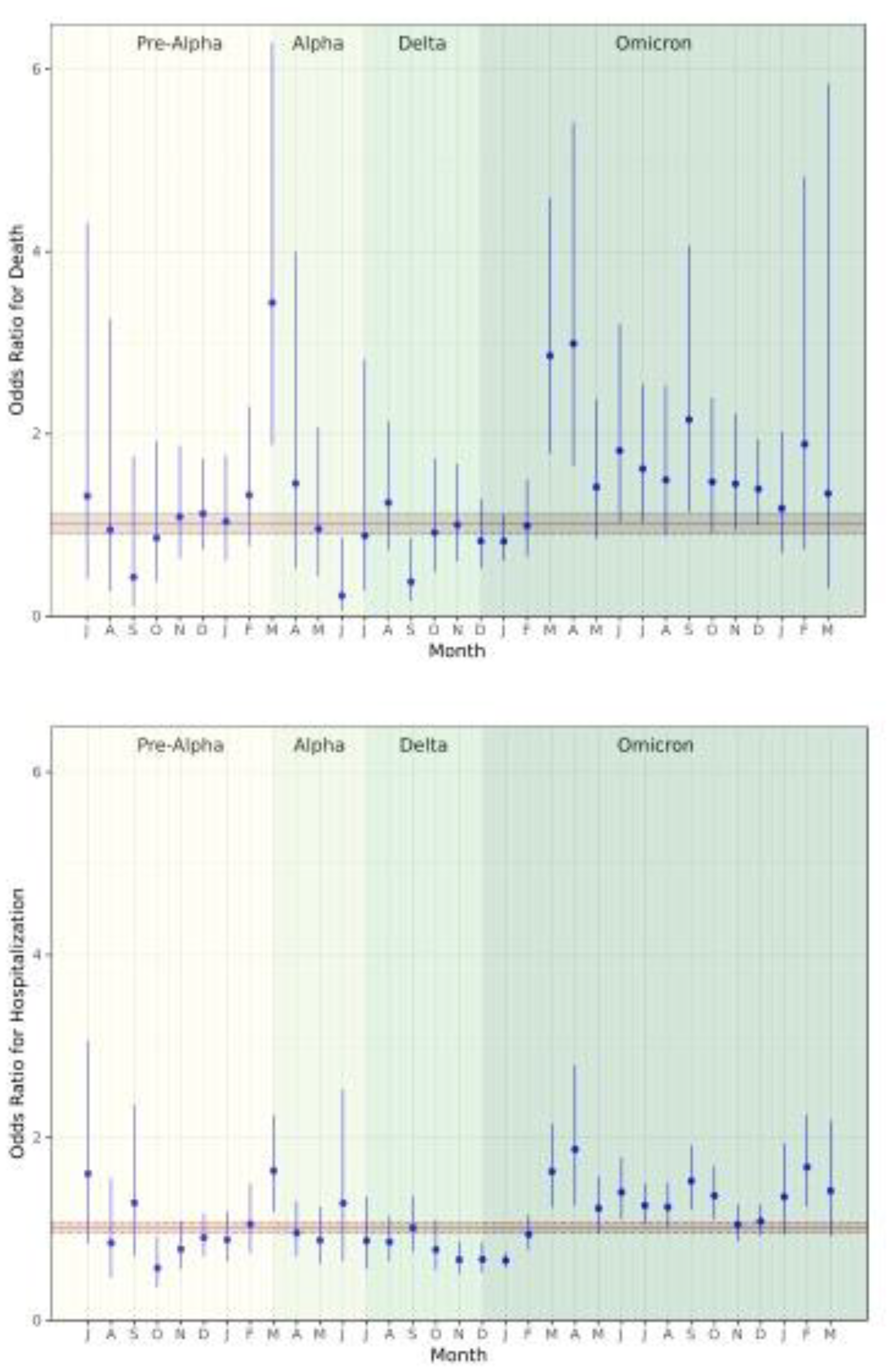
Odds ratios for death (top) and hospitalization (bottom) as binary outcomes using a logistic regression in a sensitivity analysis

Similarly, there were two months (June and September of 2021) in which prior infection reduced the risk of death, and there were seven months in which prior infection increased that risk. As in the analysis of hospitalization risk, all but one month of elevated risk took place in March 2022 or later. The ORs ranged from 0.38 (95% CI: 0.17 to 0.86) in September 2021 to 3.44 (95% CI: 1.88 to 6.29) in March 2021. The pandemic-wide OR was 1.01 (95% CI: 0.90 to 1.13). The sensitivity analyses showed very similar effects (Supplemental Material V – VIII).

### Risk of More Severe Disease

The pandemic-wide effect of prior infection on the complete cohort was slightly but significantly predictive of greater severity (Figure 4). The estimated OR of more severe disease given prior infection was 1.06 (95% confidence interval [CI]: 1.03 – 1.10).

**Figure 4:**
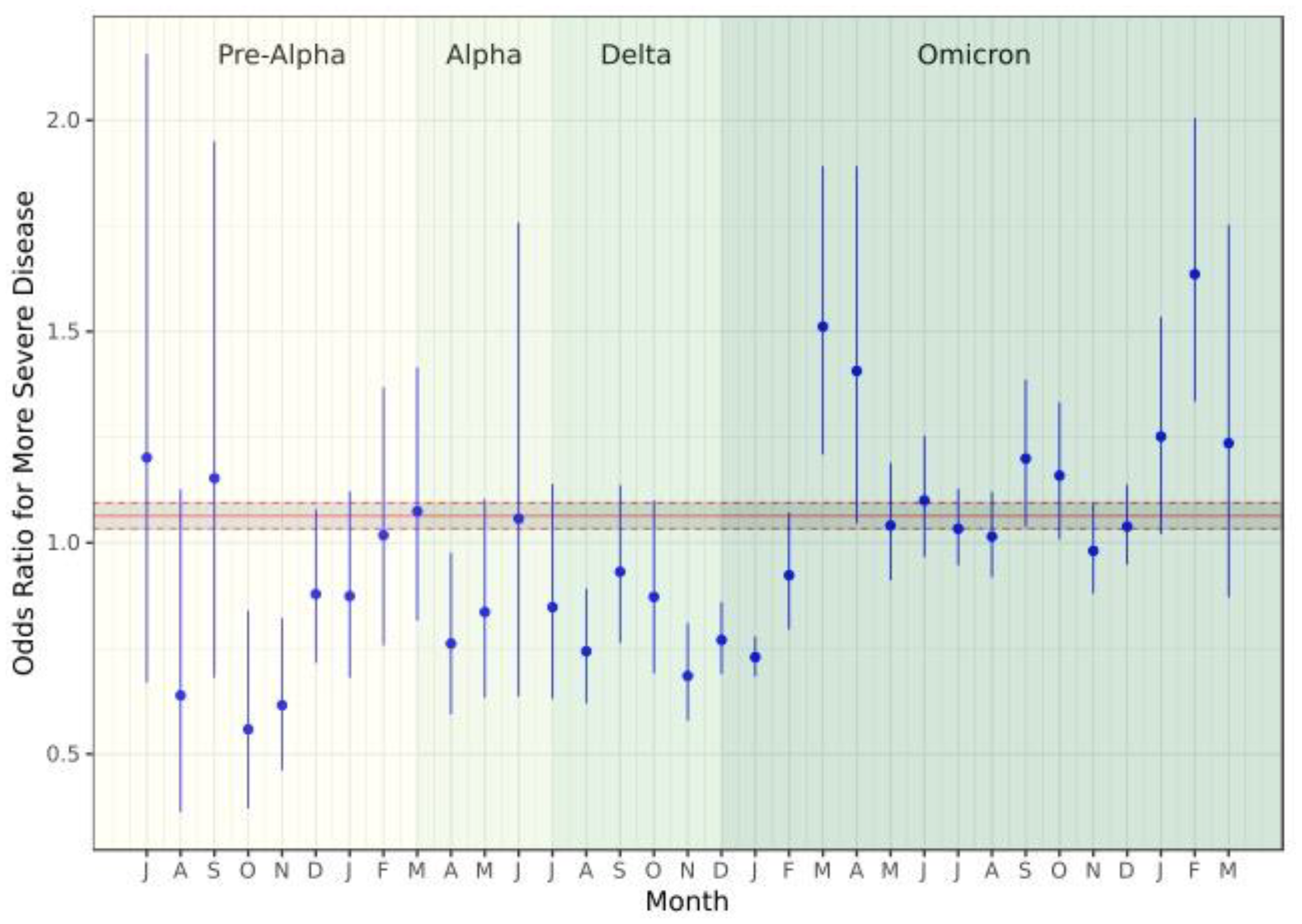
Odds ratio for more severe disease among the full cohort. The solid red line represents the point estimate of the pandemic-wide effect of prior infection, while the dashed red lines represent the 95% confidence interval around that estimate.

There was substantial heterogeneity in the impact of prior infection on COVID-19 outcomes across the pandemic, however. There was a total of 7 months where reinfection offered a protective effect, all prior to March 2022. After this point, there were 6 months in which prior infection was associated with increased odds of more severe disease. Many monthly confidence intervals were large and, in 20 of the 33 months analyzed, crossed unity.

For most of the subgroups we analyzed, prior infection had a significant association with disease severity, though there were differences in the magnitude and directionality of this effect (Supplemental Material IX). The greatest protective effect was observed among patients aged 70 or greater (OR: 0.89 [0.85 to 0.94]), although prior infection among patients in all groups aged 45 and greater was associated with a significant protective effect (OR for 45- to 59-year-old patients: 0.92 [0.87 to 0.97], OR for 60- to 69-year-old patients: 0.92 [0.86 to 0.99]). In the pandemic-wide analysis, prior infection among male patients (OR: 1.11 [1.06 to 1.16]), patients aged less than 18 years (OR: 1.34 [1.22 – 1.49]), and black / African American patients (OR: 1.08 [1.01 – 1.15]) was significantly associated with more severe disease. No subgroup had a consistently significant difference of effects compared to other groups in the monthly analysis.

The sensitivity analyses assessing the impact of our inclusion criteria and definition of second infection showed largely the same results, albeit with somewhat more attenuated effects (Supplemental Material X, XI).

## Discussion

Across the pandemic, we observed a substantial shift in how prior infection influences the risk of severe disease relative to no prior infection. Prior to March 2022, there was only one month (September 2021) in which the risks of hospitalization, death, and overall more severe disease were elevated among previously infected individuals; after that period, however, prior infection was associated with an increased risk of worse outcomes in at least half of the months included in our analysis. These findings are largely corroborated by a number of sensitivity and subgroup analyses.

The changes in the impact of prior infection that we observed must be interpreted in the context of monthly cohorts with very different characteristics. The introduction and broad uptake of vaccination, the shifting dominance of new SARS-CoV-2 variants, and the increasing time between first and second infection all likely played a role in the patterns we identified in this study. At the same time, we did not design this study to test those effects. Therefore, our interpretation of the roles played by these time-varying influences is necessarily very speculative.

Our findings may be consistent with a scenario in which infection has both protective and harmful effects of differing durations: the protective effects predominate in the short term, while harmful effects are longer lasting. In this explanation, the benefits of infection to immune readiness subside while damage to the lungs, vasculature, and other systems have not been fully healed.(11,12) Prior studies have found that COVID-19 confers protection against subsequent infection for approximately one year.(13) The point in the pandemic when average time between first and second infection began to exceed one year was the same point at which the first harmful effects of prior infection became more common. This lends some credibility to our hypothesis, as does the appearance of potential harm from previous infection at points when SARS-CoV-2 had mutated most dramatically(14) and immunologic cross-protection conferred by prior infection, especially more remote previous infection, may have been minimal or non-existent.

At least three previous studies of reinfection severity have been published with contradictory results. The studies by Abu-Raddad, et al. and Mensah, et al. used nationwide surveillance from Qatar and the United Kingdom, respectively.(15,16) Both found significantly diminished severity among individuals with prior infection. Notably, both of these publications appeared prior to April 2022. On the other hand, Bowe, et al. found a substantially increased risk of death and multiple post-acute sequelae among patients in the United States Veterans Affairs healthcare system with more than one SARS-CoV-2 infection, although their study design was compromised by immortal time bias and issues with the timing of outcome ascertainment among singly infected individuals.(17)

Our study had multiple limitations. The presumed unreliability of vaccination data at some sites meant that we had to exclude data partners for whom we could not validate the observed prevalence of vaccination against their local vaccination rates. We also had no way of validating individual-level vaccination data. Next, we had no way of validating that first and second recorded infections were, in fact, the first or second infections that patients had experienced. In practice, the cohort of infections we deemed “first” was likely a mix of first and subsequent infections with ratios that varied across the pandemic. Although we attempted to control for this possibility by using time in the ordinal logistic regression, we may underestimate any protective effect of prior infection. Similarly, our IPW approach could not include all key variables related to risk of prior infection, such as masking and distancing behavior, given the limitations of the data set. In addition, the widespread use of home testing late in the pandemic may have led to missed mild Omicron infections, inflating the apparent disease severity for both first and second infection cohorts during this time. Finally, we cannot precisely attribute death to COVID since it was based on discharge status instead of cause of death.

## Conclusion

Our analysis of a large, diverse collection of electronic health records for COVID-19 patients revealed that prior infection produced variable effects throughout the pandemic: in its first two years, prior infection was largely protective to neutral, but after March 2022, prior infection raised the risk of more severe outcomes relative to patients without prior infection. Our results imply that acquired immunity through infection cannot be relied on to provide comprehensive protection against severe disease. In turn, public health efforts should benefit from continued investments in vaccination and treatment.

## Data Availability

Data are available online through the National COVID Cohort Collaborative (https://ncats.nih.gov/n3c) with signed data use agreement and project approval.

## Acknowledgment

The analyses described in this publication were conducted with data or tools accessed through the NCATS N3C Data Enclave https://covid.cd2h.org and N3C Attribution & Publication Policy v 1.2-2020-08-25b supported by NCATS U24 TR002306, Axle Informatics Subcontract: NCATS-P00438-B. This research was possible because of the patients whose information is included within the data and the organizations (https://ncats.nih.gov/n3c/resources/data-contribution/data-transfer-agreement-signatories) and scientists who have contributed to the on-going development of this community resource [https://doi.org/10.1093/jamia/ocaa196].

