## Supplemental material for "Influence of Prior SARS-CoV-2 Infection on COVID-19 Severity: Evidence from the National COVID Cohort Collaborative"

### I. Comorbidities used in inverse probability of treatment weight calculations

Tuberculosis, mild liver disease, moderate-to-severe liver disease, thalassemia, rheumatological disease, dementia, congestive heart failure, substance use disorder, Down syndrome, kidney disease, malignant cancer, uncomplicated diabetes, complicated diabetes, cerebrovascular disease, peripheral vascular disease, pregnancy, hemiplegia / paraplegia, psychosis, obesity, coronary artery disease, systemic corticosteroids, depression, metastatic solid tumor cancers, HIV infection, chronic lung disease, sickle cell disease, myocardial infarction, cardiomyopathies, hypertension, other immunocompromised status, tobacco smoking, and solid organ / blood stem cell transplant.

II. Love plots of covariate balance

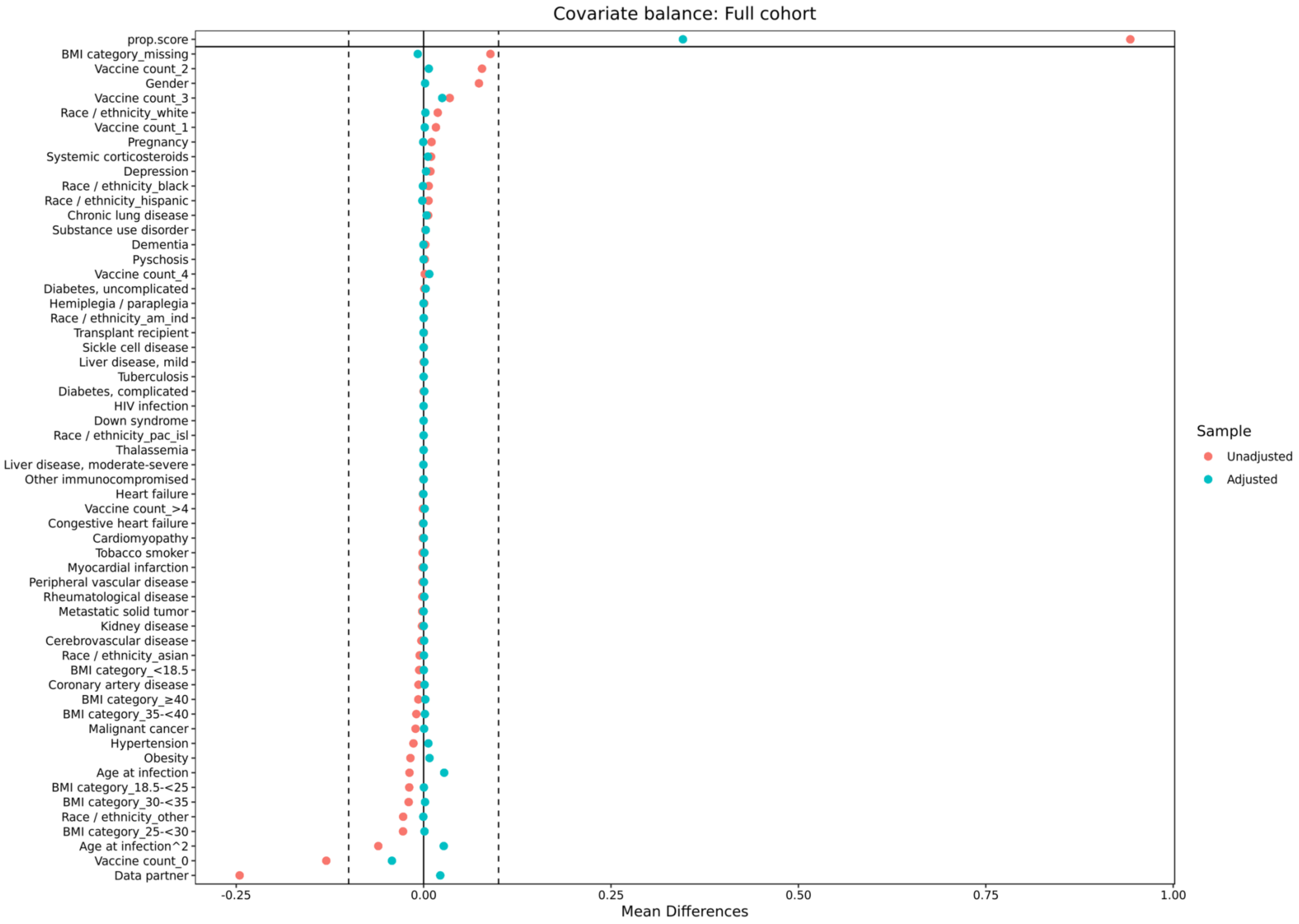

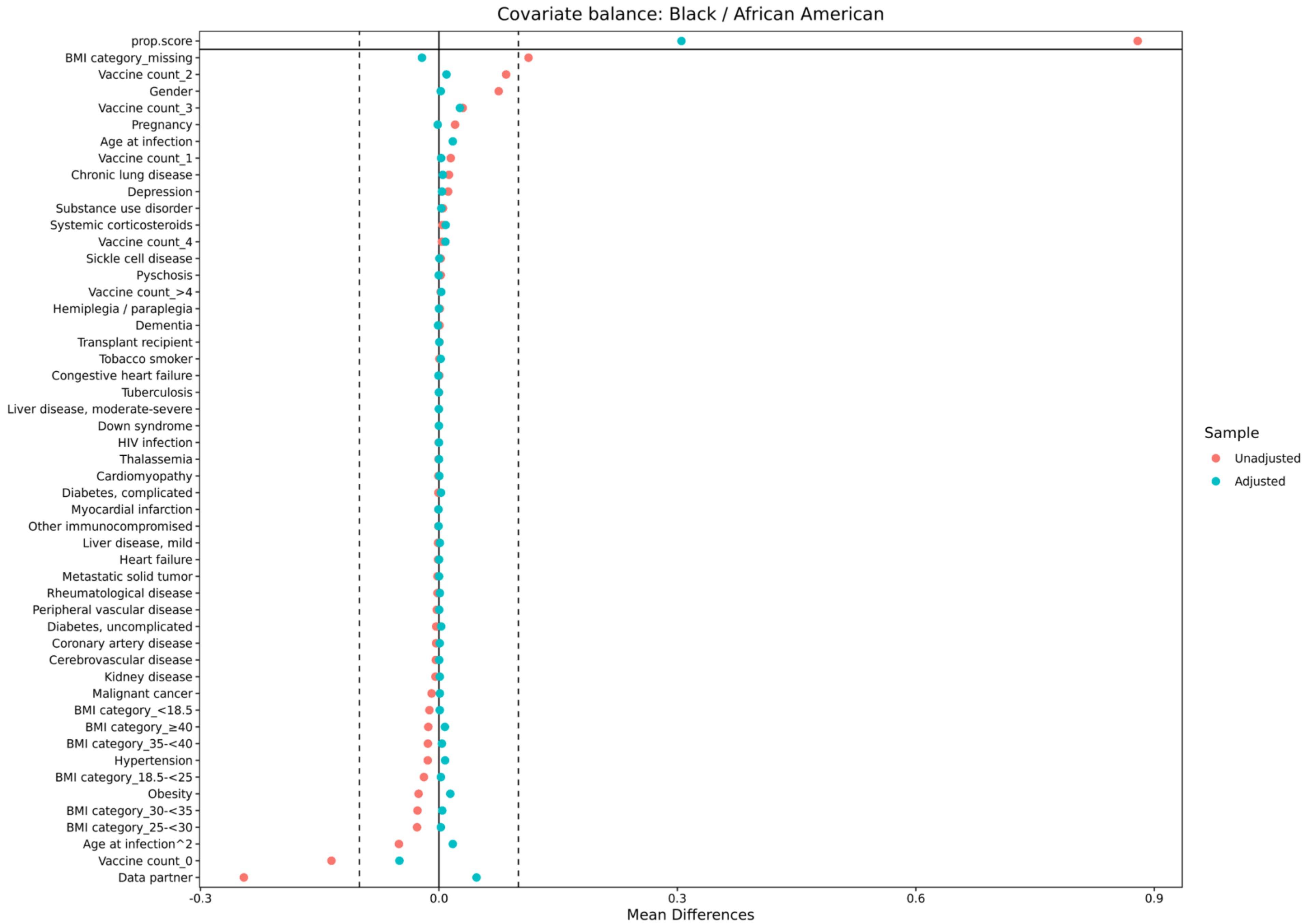
Covariate balance: Black / African American
prop.score
BMI category_missing
Vaccine count_2
Gender
Vaccine count_3
Pregnancy
Age at infection
Vaccine count_1
Chronic lung disease
Depression
Substance use disorder
Systemic corticosteroids
Vaccine count_4
Sickle cell disease
Pyschosis
Vaccine count_>4
Hemiplegia / paraplegia
Dementia
Transplant recipient
Tobacco smoker
Congestive heart failure
Tuberculosis
Liver disease, moderate-severe
Down syndrome
HIV infection
Thalassemia
Cardiomyopathy
Diabetes, complicated
Myocardial infarction
Other immunocompromised
Liver disease, mild
Heart failure
Metastatic solid tumor
Rheumatological disease
Peripheral vascular disease
Diabetes, uncomplicated
Coronary artery disease
Cerebrovascular disease
Kidney disease
Malignant cancer
BMI category_<18.5
BMI category_≥40
BMI category_35-<40
Hypertension
BMI category_18.5-<25
Obesity
BMI category_30-<35
BMI category_25-<30
Age at infection^2
Vaccine count_0
Data partner
-0.3
0.0
0.3
0.6
0.9
Mean Differences
Sample
Unadjusted
Adjusted

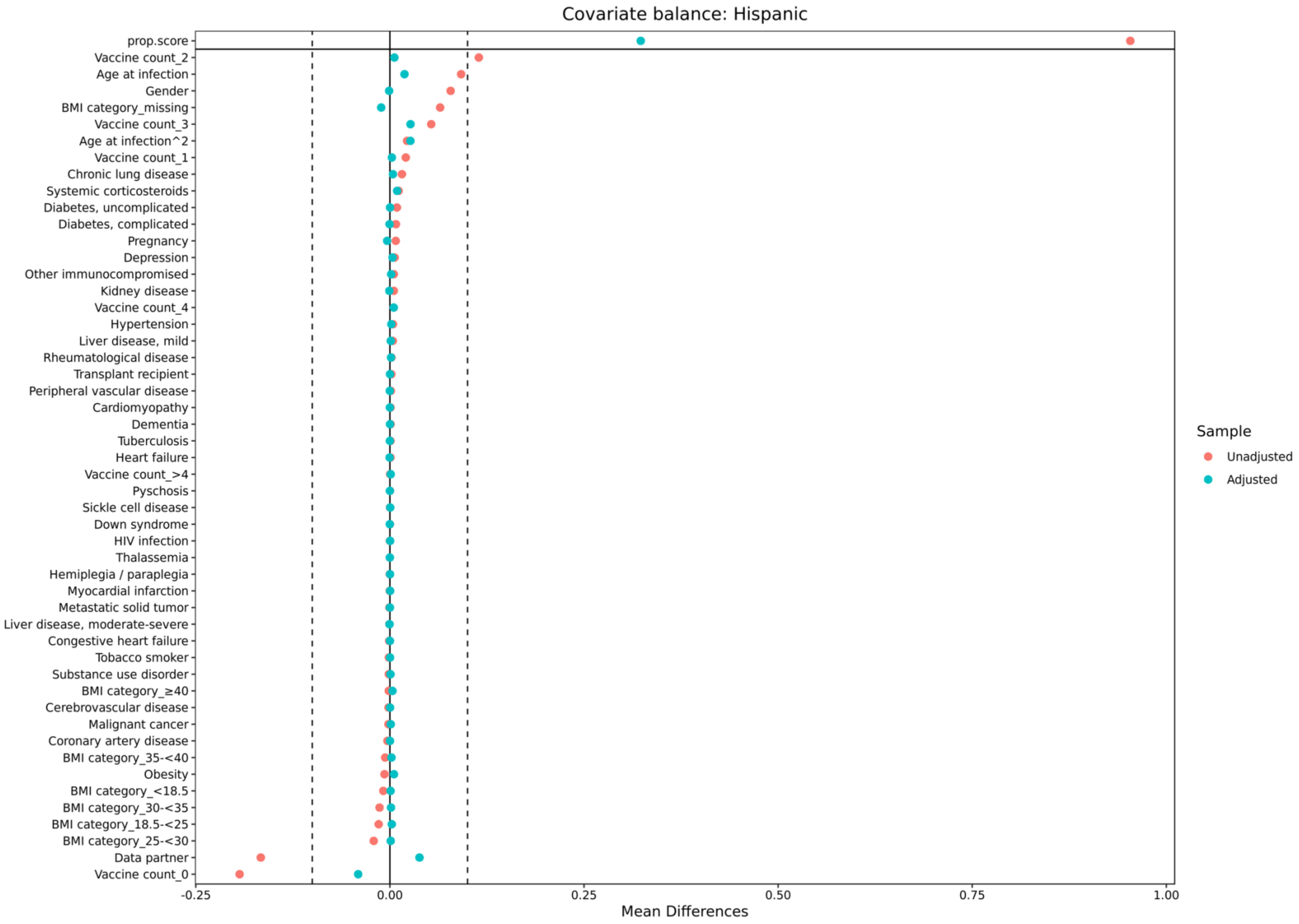

Covariate balance: Hispanic
prop.score
Vaccine count_2
Age at infection
Gender
BMI category_missing
Vaccine count_3
Age at infection^2
Vaccine count_1
Chronic lung disease
Systemic corticosteroids
Diabetes, uncomplicated
Diabetes, complicated
Pregnancy
Depression
Other immunocompromised
Kidney disease
Vaccine count_4
Hypertension
Liver disease, mild
Rheumatological disease
Transplant recipient
Peripheral vascular disease
Cardiomyopathy
Dementia
Tuberculosis
Heart failure
Vaccine count_>4
Pyschosis
Sickle cell disease
Down syndrome
HIV infection
Thalassemia
Hemiplegia / paraplegia
Myocardial infarction
Metastatic solid tumor
Liver disease, moderate-severe
Congestive heart failure
Tobacco smoker
Substance use disorder
BMI category_≥40
Cerebrovascular disease
Malignant cancer
Coronary artery disease
BMI category_35-<40
Obesity
BMI category_<18.5
BMI category_30-<35
BMI category_18.5-<25
BMI category_25-<30
Data partner
Vaccine count_0
-0.25
0.00
0.25
0.50
0.75
1.00
Mean Differences
Sample
Unadjusted
Adjusted

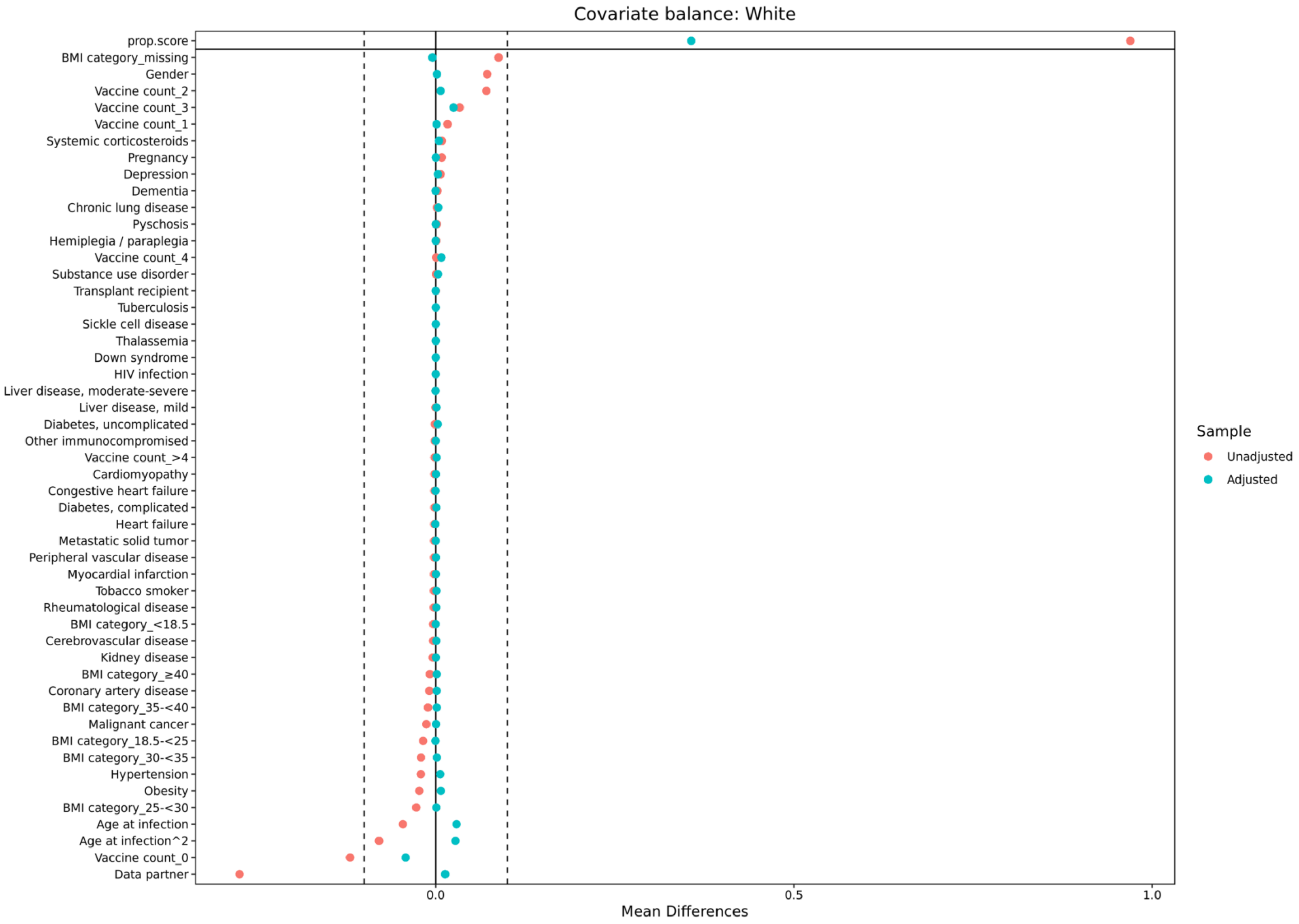
Covariate balance: White
prop.score
BMI category_missing
Gender
Vaccine count_2
Vaccine count_3
Vaccine count_1
Systemic corticosteroids
Pregnancy
Depression
Dementia
Chronic lung disease
Pyschosis
Hemiplegia / paraplegia
Vaccine count_4
Substance use disorder
Transplant recipient
Tuberculosis
Sickle cell disease
Thalassemia
Down syndrome
HIV infection
Liver disease, moderate-severe
Liver disease, mild
Diabetes, uncomplicated
Other immunocompromised
Vaccine count_>4
Cardiomyopathy
Congestive heart failure
Diabetes, complicated
Heart failure
Metastatic solid tumor
Peripheral vascular disease
Myocardial infarction
Tobacco smoker
Rheumatological disease
BMI category_<18.5
Cerebrovascular disease
Kidney disease
BMI category_≥40
Coronary artery disease
BMI category_35-<40
Malignant cancer
BMI category_18.5-<25
BMI category_30-<35
Hypertension
Obesity
BMI category_25-<30
Age at infection
Age at infection^2
Vaccine count_0
Data partner
0.0
0.5
1.0
Mean Differences
Sample
Unadjusted
Adjusted

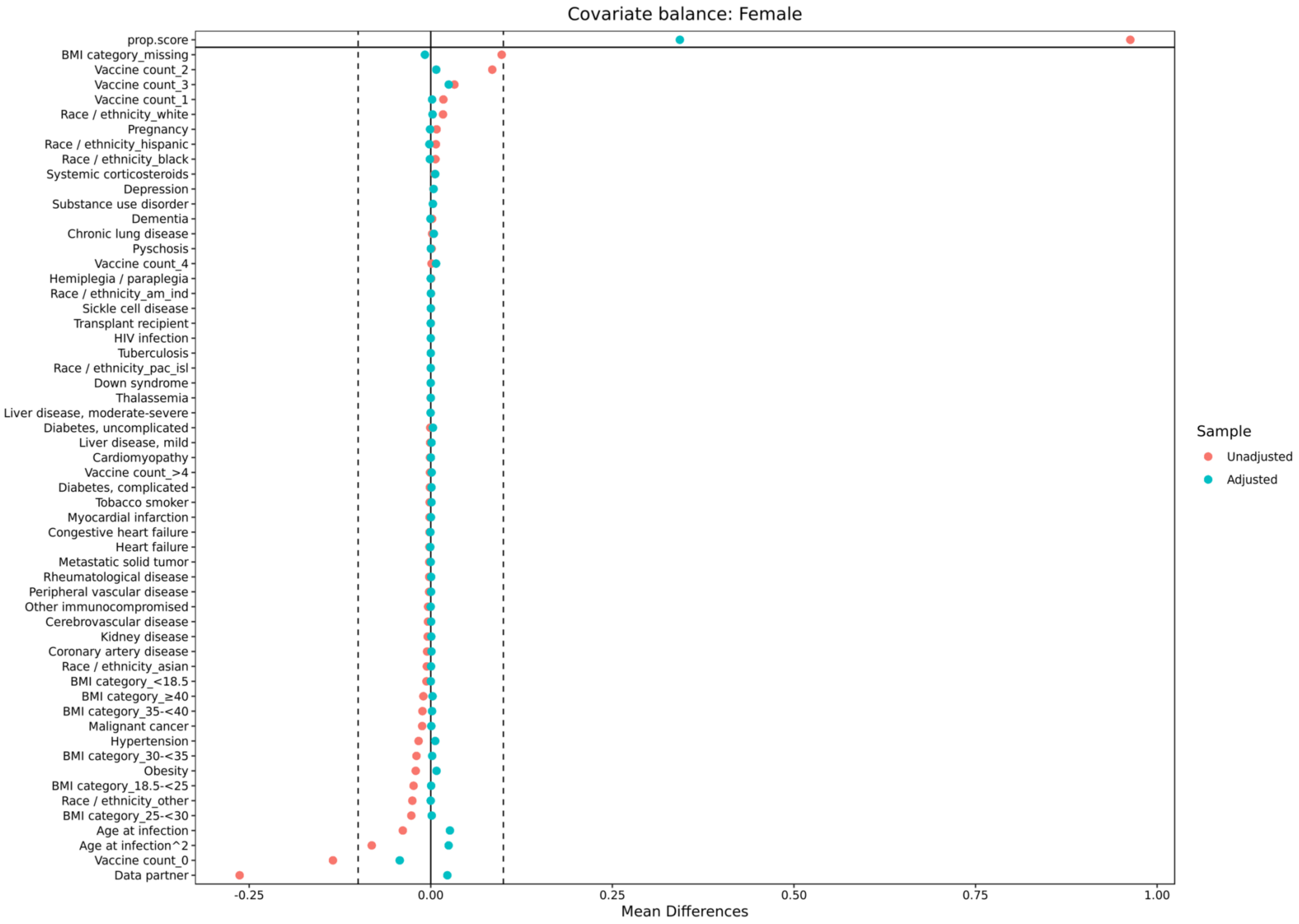
Covariate balance: Female
prop.score
BMI category_missing
Vaccine count_2
Vaccine count_3
Vaccine count_1
Race / ethnicity_white
Pregnancy
Race / ethnicity_hispanic
Race / ethnicity_black
Systemic corticosteroids
Depression
Substance use disorder
Dementia
Chronic lung disease
Pyschosis
Vaccine count_4
Hemiplegia / paraplegia
Race / ethnicity_am_ind
Sickle cell disease
Transplant recipient
HIV infection
Tuberculosis
Race / ethnicity_pac_isl
Down syndrome
Thalassemia
Liver disease, moderate-severe
Diabetes, uncomplicated
Liver disease, mild
Cardiomyopathy
Vaccine count_>4
Diabetes, complicated
Tobacco smoker
Myocardial infarction
Congestive heart failure
Heart failure
Metastatic solid tumor
Rheumatological disease
Peripheral vascular disease
Other immunocompromised
Cerebrovascular disease
Kidney disease
Coronary artery disease
Race / ethnicity_asian
BMI category_<18.5
BMI category_≥40
BMI category_35-<40
Malignant cancer
Hypertension
BMI category_30-<35
Obesity
BMI category_18.5-<25
Race / ethnicity_other
BMI category_25-<30
Age at infection
Age at infection^2
Vaccine count_0
Data partner
-0.25
0.00
0.25
0.50
0.75
1.00
Mean Differences
Sample
Unadjusted
Adjusted

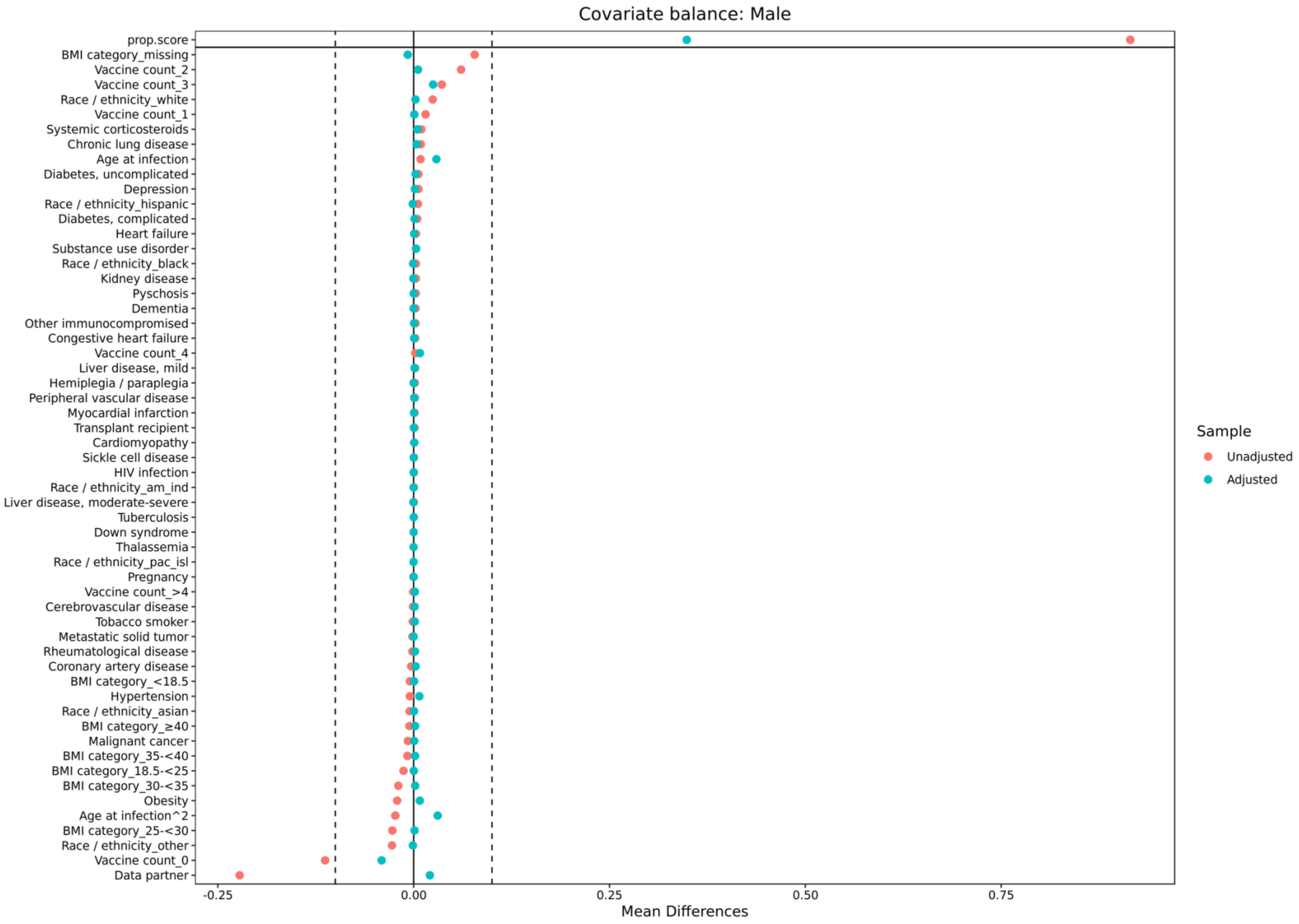

Covariate balance: Male
prop.score
BMI category_missing
Vaccine count_2
Vaccine count_3
Race / ethnicity_white
Vaccine count_1
Systemic corticosteroids
Chronic lung disease
Age at infection
Diabetes, uncomplicated
Depression
Race / ethnicity_hispanic
Diabetes, complicated
Heart failure
Substance use disorder
Race / ethnicity_black
Kidney disease
Pyschosis
Dementia
Other immunocompromised
Congestive heart failure
Vaccine count_4
Liver disease, mild
Hemiplegia / paraplegia
Peripheral vascular disease
Myocardial infarction
Transplant recipient
Cardiomyopathy
Sickle cell disease
HIV infection
Race / ethnicity_am_ind
Liver disease, moderate-severe
Tuberculosis
Down syndrome
Thalassemia
Race / ethnicity_pac_isl
Pregnancy
Vaccine count_>4
Cerebrovascular disease
Tobacco smoker
Metastatic solid tumor
Rheumatological disease
Coronary artery disease
BMI category_<18.5
Hypertension
Race / ethnicity_asian
BMI category_≥40
Malignant cancer
BMI category_35-<40
BMI category_18.5-<25
BMI category_30-<35
Obesity
Age at infection^2
BMI category_25-<30
Race / ethnicity_other
Vaccine count_0
Data partner
-0.25
0.00
0.25
0.50
0.75
Mean Differences
Sample
Unadjusted
Adjusted

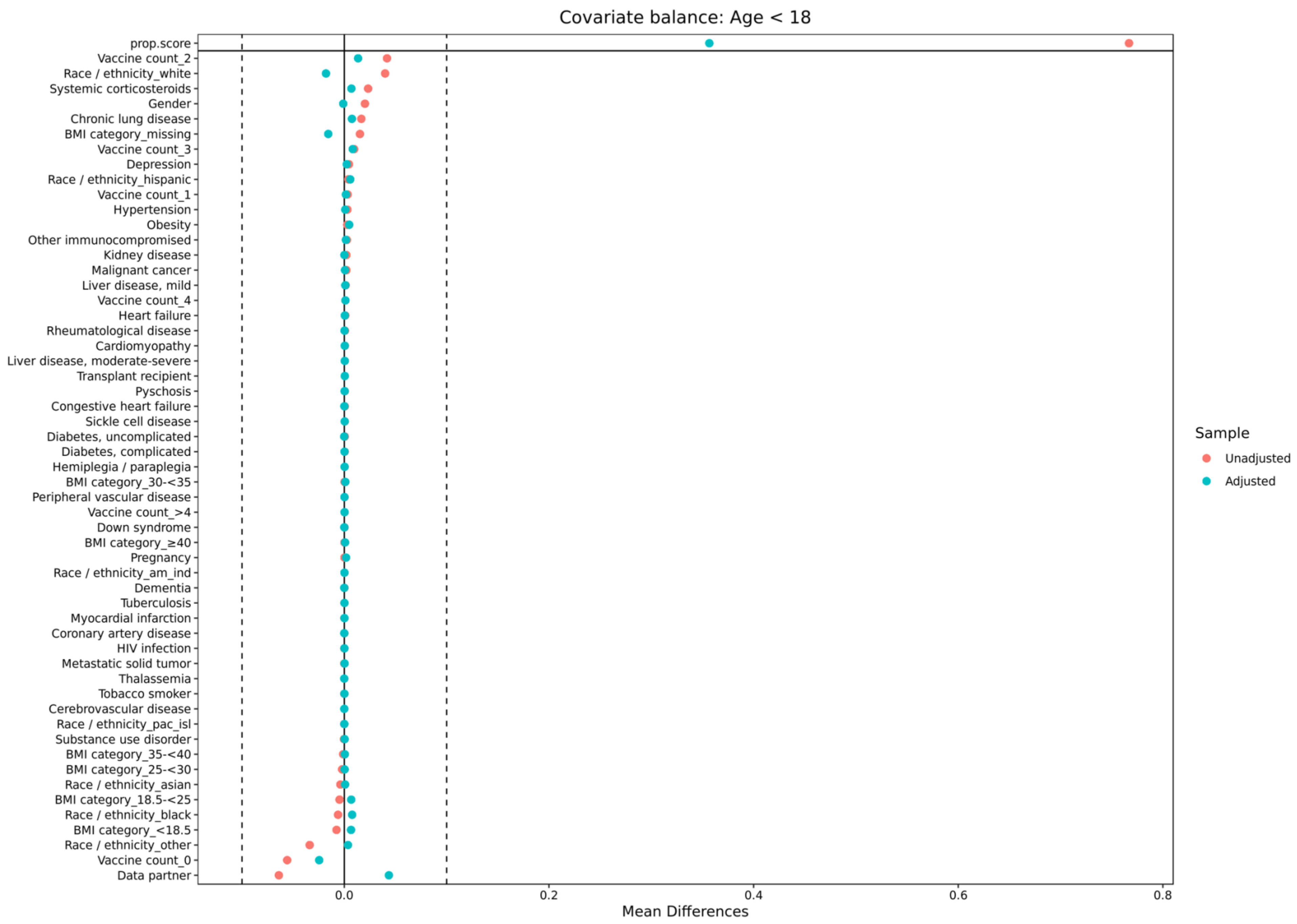
Covariate balance: Age < 18
prop.score
Vaccine count_2
Race / ethnicity_white
Systemic corticosteroids
Gender
Chronic lung disease
BMI category_missing
Vaccine count_3
Depression
Race / ethnicity_hispanic
Vaccine count_1
Hypertension
Obesity
Other immunocompromised
Kidney disease
Malignant cancer
Liver disease, mild
Vaccine count_4
Heart failure
Rheumatological disease
Cardiomyopathy
Liver disease, moderate-severe
Transplant recipient
Pyschosis
Congestive heart failure
Sickle cell disease
Diabetes, uncomplicated
Diabetes, complicated
Hemiplegia / paraplegia
BMI category_30-<35
Peripheral vascular disease
Vaccine count_>4
Down syndrome
BMI category_≥40
Pregnancy
Race / ethnicity_am_ind
Dementia
Tuberculosis
Myocardial infarction
Coronary artery disease
HIV infection
Metastatic solid tumor
Thalassemia
Tobacco smoker
Cerebrovascular disease
Race / ethnicity_pac_isl
Substance use disorder
BMI category_35-<40
BMI category_25-<30
Race / ethnicity_asian
BMI category_18.5-<25
Race / ethnicity_black
BMI category_<18.5
Race / ethnicity_other
Vaccine count_0
Data partner
0.0
0.2
0.4
0.6
0.8
Mean Differences
Sample
Unadjusted
Adjusted

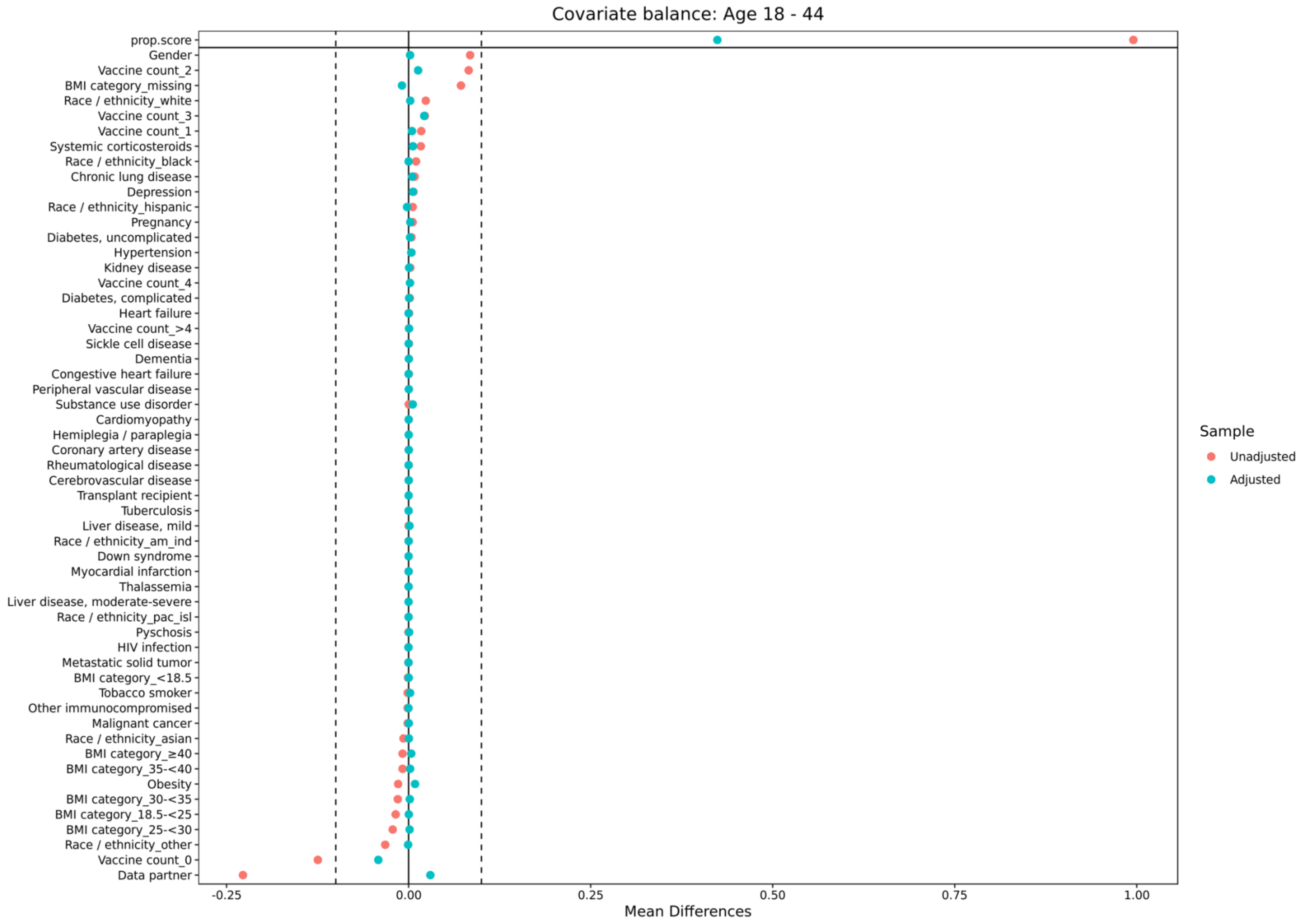

Covariate balance: Age 18 - 44
prop.score
Gender
Vaccine count_2
BMI category_missing
Race / ethnicity_white
Vaccine count_3
Vaccine count_1
Systemic corticosteroids
Race / ethnicity_black
Chronic lung disease
Depression
Race / ethnicity_hispanic
Pregnancy
Diabetes, uncomplicated
Hypertension
Kidney disease
Vaccine count_4
Diabetes, complicated
Heart failure
Vaccine count_>4
Sickle cell disease
Dementia
Congestive heart failure
Peripheral vascular disease
Substance use disorder
Cardiomyopathy
Hemiplegia / paraplegia
Coronary artery disease
Rheumatological disease
Cerebrovascular disease
Transplant recipient
Tuberculosis
Liver disease, mild
Race / ethnicity_am_ind
Down syndrome
Myocardial infarction
Thalassemia
Liver disease, moderate-severe
Race / ethnicity_pac_isl
Pyschosis
HIV infection
Metastatic solid tumor
BMI category_<18.5
Tobacco smoker
Other immunocompromised
Malignant cancer
Race / ethnicity_asian
BMI category_≥40
BMI category_35-<40
Obesity
BMI category_30-<35
BMI category_18.5-<25
BMI category_25-<30
Race / ethnicity_other
Vaccine count_0
Data partner
-0.25
0.00
0.25
0.50
0.75
1.00
Mean Differences
Sample
Unadjusted
Adjusted

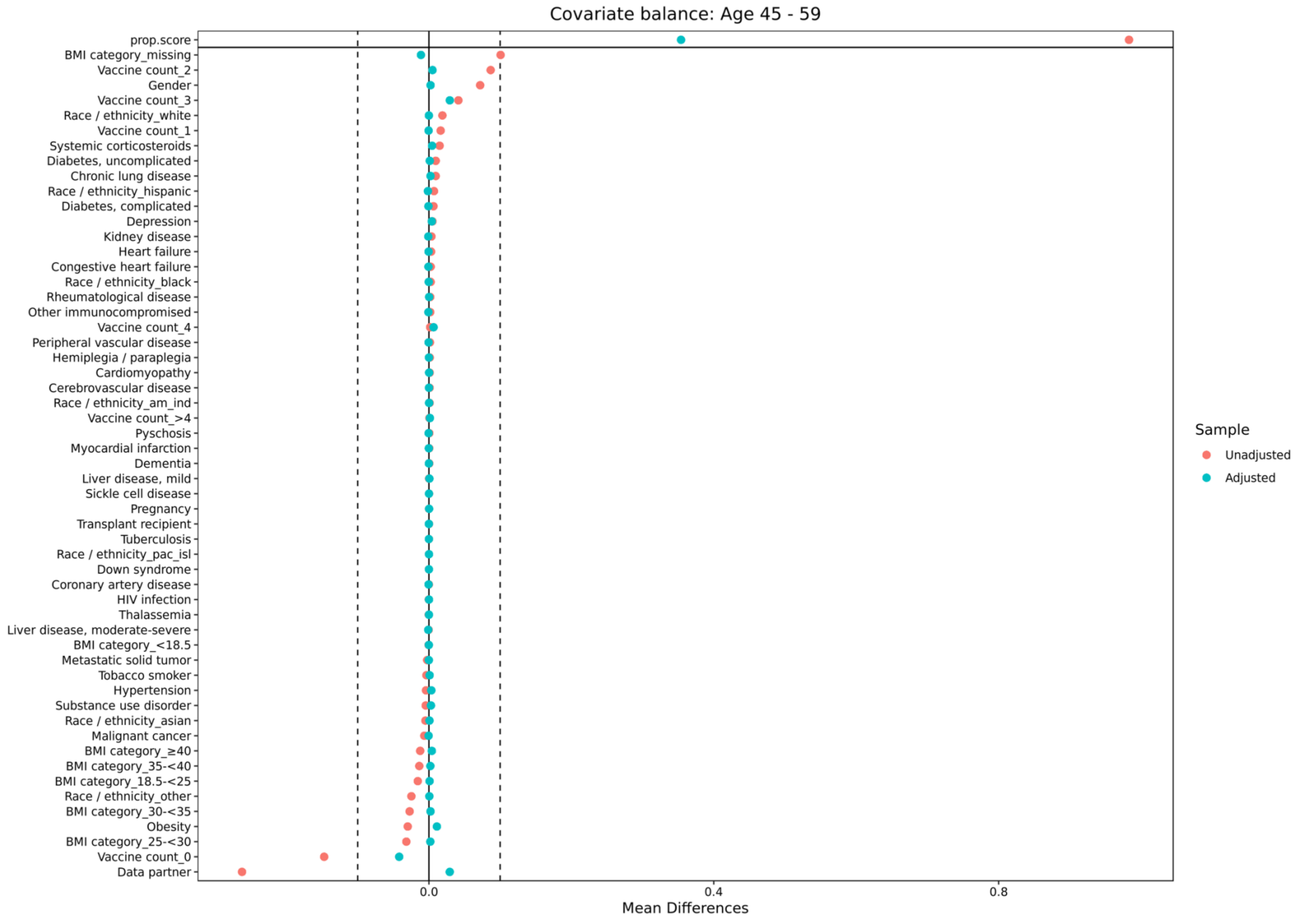

Covariate balance: Age 45 - 59
prop.score
BMI category_missing
Vaccine count_2
Gender
Vaccine count_3
Race / ethnicity_white
Vaccine count_1
Systemic corticosteroids
Diabetes, uncomplicated
Chronic lung disease
Race / ethnicity_hispanic
Diabetes, complicated
Depression
Kidney disease
Heart failure
Congestive heart failure
Race / ethnicity_black
Rheumatological disease
Other immunocompromised
Vaccine count_4
Peripheral vascular disease
Hemiplegia / paraplegia
Cardiomyopathy
Cerebrovascular disease
Race / ethnicity_am_ind
Vaccine count_>4
Pyschosis
Myocardial infarction
Dementia
Liver disease, mild
Sickle cell disease
Pregnancy
Transplant recipient
Tuberculosis
Race / ethnicity_pac_isl
Down syndrome
Coronary artery disease
HIV infection
Thalassemia
Liver disease, moderate-severe
BMI category_<18.5
Metastatic solid tumor
Tobacco smoker
Hypertension
Substance use disorder
Race / ethnicity_asian
Malignant cancer
BMI category_≥40
BMI category_35-<40
BMI category_18.5-<25
Race / ethnicity_other
BMI category_30-<35
Obesity
BMI category_25-<30
Vaccine count_0
Data partner
0.0
0.4
0.8
Mean Differences
Sample
Unadjusted
Adjusted

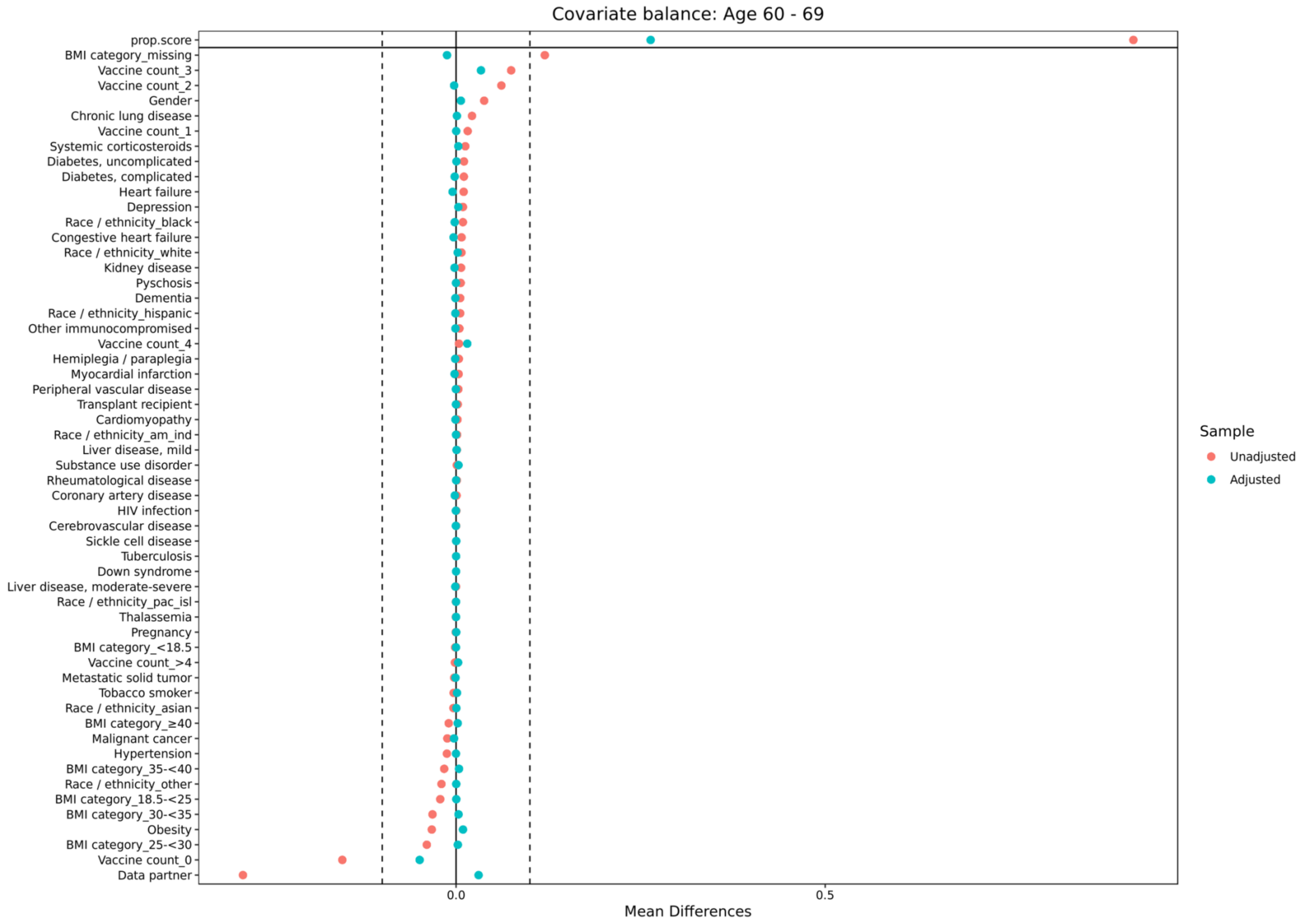
Covariate balance: Age 60 - 69
prop.score
BMI category_missing
Vaccine count_3
Vaccine count_2
Gender
Chronic lung disease
Vaccine count_1
Systemic corticosteroids
Diabetes, uncomplicated
Diabetes, complicated
Heart failure
Depression
Race / ethnicity_black
Congestive heart failure
Race / ethnicity_white
Kidney disease
Pyschosis
Dementia
Race / ethnicity_hispanic
Other immunocompromised
Vaccine count_4
Hemiplegia / paraplegia
Myocardial infarction
Peripheral vascular disease
Transplant recipient
Cardiomyopathy
Race / ethnicity_am_ind
Liver disease, mild
Substance use disorder
Rheumatological disease
Coronary artery disease
HIV infection
Cerebrovascular disease
Sickle cell disease
Tuberculosis
Down syndrome
Liver disease, moderate-severe
Race / ethnicity_pac_isl
Thalassemia
Pregnancy
BMI category_<18.5
Vaccine count_>4
Metastatic solid tumor
Tobacco smoker
Race / ethnicity_asian
BMI category_≥40
Malignant cancer
Hypertension
BMI category_35-<40
Race / ethnicity_other
BMI category_18.5-<25
BMI category_30-<35
Obesity
BMI category_25-<30
Vaccine count_0
Data partner
0.0
0.5
Mean Differences
Sample
Unadjusted
Adjusted

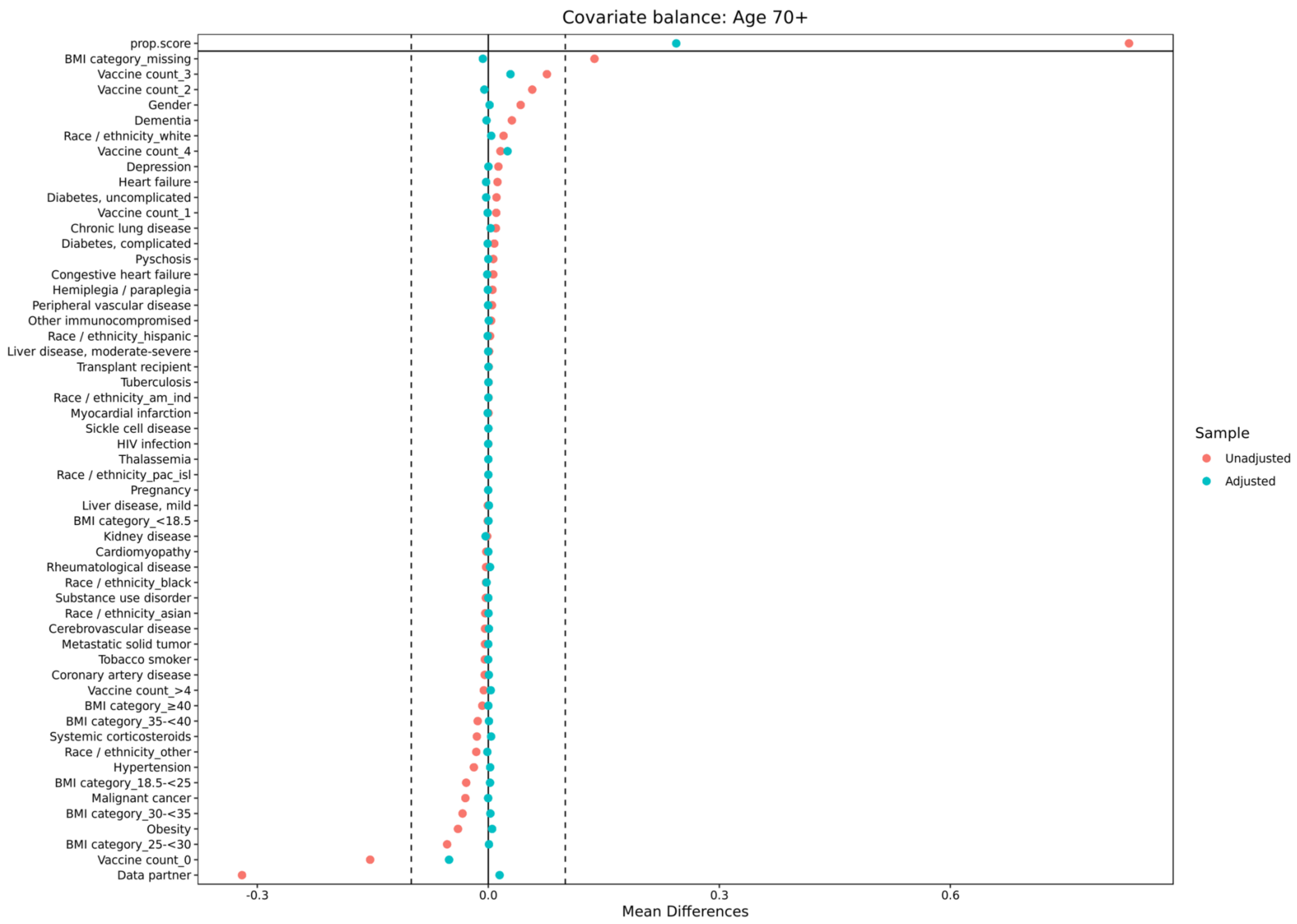
Covariate balance: Age 70+
prop.score
BMI category_missing
Vaccine count_3
Vaccine count_2
Gender
Dementia
Race / ethnicity_white
Vaccine count_4
Depression
Heart failure
Diabetes, uncomplicated
Vaccine count_1
Chronic lung disease
Diabetes, complicated
Pyschosis
Congestive heart failure
Hemiplegia / paraplegia
Peripheral vascular disease
Other immunocompromised
Race / ethnicity_hispanic
Liver disease, moderate-severe
Transplant recipient
Tuberculosis
Race / ethnicity_am_ind
Myocardial infarction
Sickle cell disease
HIV infection
Thalassemia
Race / ethnicity_pac_isl
Pregnancy
Liver disease, mild
BMI category_<18.5
Kidney disease
Cardiomyopathy
Rheumatological disease
Race / ethnicity_black
Substance use disorder
Race / ethnicity_asian
Cerebrovascular disease
Metastatic solid tumor
Tobacco smoker
Coronary artery disease
Vaccine count_>4
BMI category_≥40
BMI category_35-<40
Systemic corticosteroids
Race / ethnicity_other
Hypertension
BMI category_18.5-<25
Malignant cancer
BMI category_30-<35
Obesity
BMI category_25-<30
Vaccine count_0
Data partner
-0.3
0.0
0.3
0.6
Mean Differences
Sample
Unadjusted
Adjusted

III. Count of patients by infection status in each month

| Month | First Infections | Second Infections |
|---|---|---|
| July-20 | 27,095 | 166 |
| August-20 | 29,635 | 191 |
| September-20 | 28,622 | 209 |
| October-20 | 61,807 | 363 |
| November-20 | 153,211 | 938 |
| December-20 | 159,790 | 1,231 |
| January-21 | 106,250 | 1,164 |
| February-21 | 35,763 | 830 |
| March-21 | 28,889 | 885 |
| April-21 | 34,506 | 901 |
| May-21 | 21,196 | 647 |
| June-21 | 8,502 | 350 |
| July-21 | 22,421 | 575 |
| August-21 | 96,179 | 2,003 |
| September-21 | 97,176 | 1,339 |
| October-21 | 54,358 | 1,218 |
| November-21 | 80,685 | 2,249 |
| December-21 | 171,394 | 10,707 |
| January-22 | 351,754 | 40,905 |
| February-22 | 47,466 | 5,306 |
| March-22 | 10,953 | 1,341 |
| April-22 | 18,708 | 1,768 |
| May-22 | 51,186 | 5,545 |
| June-22 | 53,394 | 7,603 |
| July-22 | 68,884 | 12,553 |
| August-22 | 66,151 | 13,697 |
| September-22 | 32,466 | 5,717 |
| October-22 | 23,418 | 4,479 |
| November-22 | 28,256 | 7,087 |
| December-22 | 42,834 | 9,531 |
| January-23 | 21,630 | 4,395 |
| February-23 | 10,403 | 1,094 |
| March-23 | 3,491 | 404 |

IV. Distribution of covariates across first and second infections

| Covariate | First infection (%) | Second infection (%) |
|---|---|---|
| Health system: A | 9.3% | 6.1% |
| Health system: B | 73.7% | 83.9% |
| Health system: C | 5.1% | 3.4% |
| Health system: D | 2.7% | 1.6% |
| Health system: E | 2.4% | 2.5% |
| Health system: F | 1.8% | 1.0% |
| Health system: G | 4.4% | 1.2% |
| Health system: H | 0.6% | 0.3% |
| Missing postal code | 1.1% | 0.4% |
| Vaccine doses: 0 | 68.4% | 55.0% |
| Vaccine doses: 1 | 3.6% | 5.3% |
| Vaccine doses: 2 | 16.3% | 24.3% |
| Vaccine doses: 3 | 9.2% | 12.8% |
| Vaccine doses: 4 | 1.9% | 2.1% |
| Vaccine doses: >4 | 0.6% | 0.5% |
| BMI: <18.5 | 1.5% | 0.9% |
| BMI: 18.5 – 24.9 | 4.4% | 2.5% |
| BMI: 25 – 29.9 | 6.2% | 3.4% |
| BMI: 30 – 34.9 | 5.3% | 3.2% |
| BMI: 35 – 40 | 3.2% | 2.2% |
| BMI: >40 | 3.4% | 2.6% |
| BMI: Missing | 76.0% | 85.1% |
| Tuberculosis | 0.05% | 0.05% |
| Liver disease, mild | 2.8% | 2.8% |
| Liver disease, moderate/severe | 0.4% | 0.4% |
| Thalassemia | 0.05% | 0.03% |
| Rheumatological disease | 3.3% | 3.1% |
| Dementia | 1.3% | 1.5% |
| Congestive heart failure | 2.5% | 2.4% |
| Substance use disorder | 6.9% | 7.2% |
| Down syndrome | 0.01% | <0.01% |
| Kidney disease | 5.0% | 4.7% |
| Malignant cancer | 4.7% | 3.6% |
| Diabetes, uncomplicated | 9.8% | 9.8% |
| Diabetes, complicated | 4.8% | 4.8% |
| Cerebrovascular disease | 2.2% | 1.9% |
| Peripheral vascular disease | 2.3% | 2.2% |
| Pregnancy | 4.6% | 5.7% |
| Hemiplegia / paraplegia | 0.5% | 0.6% |
| Psychosis | 0.6% | 0.8% |
| Obesity | 17.8% | 16.0% |
| Coronary artery disease | 4.9% | 4.2% |
| Systemic corticosteroids | 19.9% | 20.9% |
| Depression | 9.5% | 10.4% |
| Metastatic solid tumor cancers | 0.6% | 0.4% |
| HIV infection | 0.2% | 0.2% |
| Chronic lung disease | 10.0% | 10.6% |
| Sickle cell disease | 0.1% | 0.1% |
| Myocardial infarction | 1.8% | 1.6% |
| Cardiomyopathies | 1.5% | 1.3% |
| Hypertension | 21.5% | 20.0% |
| Other immunocompromised | 3.9% | 3.9% |

| | | |
|---|---|---|
| Tobacco smoking | 2.1% | 1.9% |
| Solid organ / blood stem cell transplant | 0.2% | 0.2% |

V. Risk of hospitalization: No lookback period

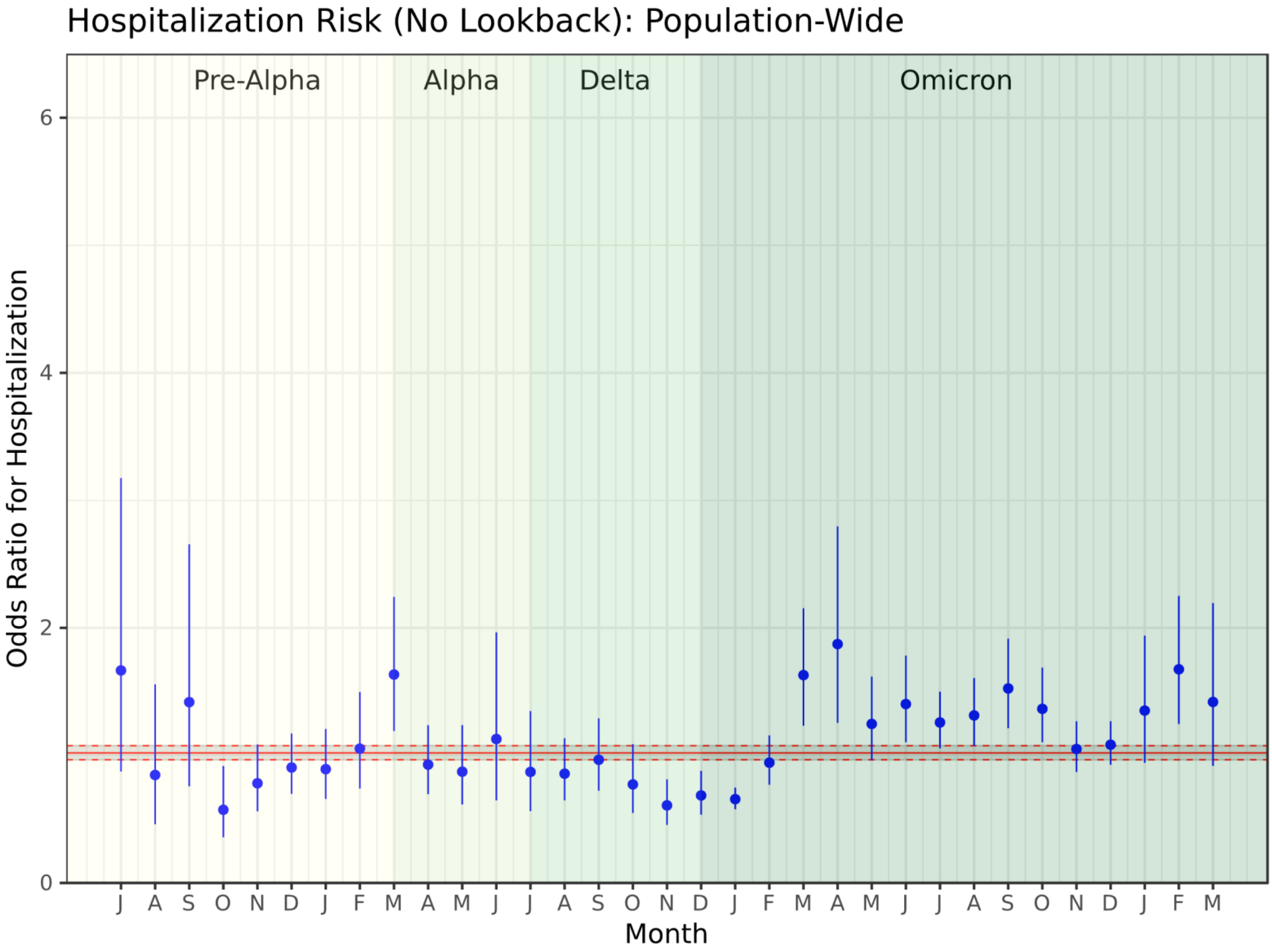

VI. Risk of hospitalization: 90-day reinfection window

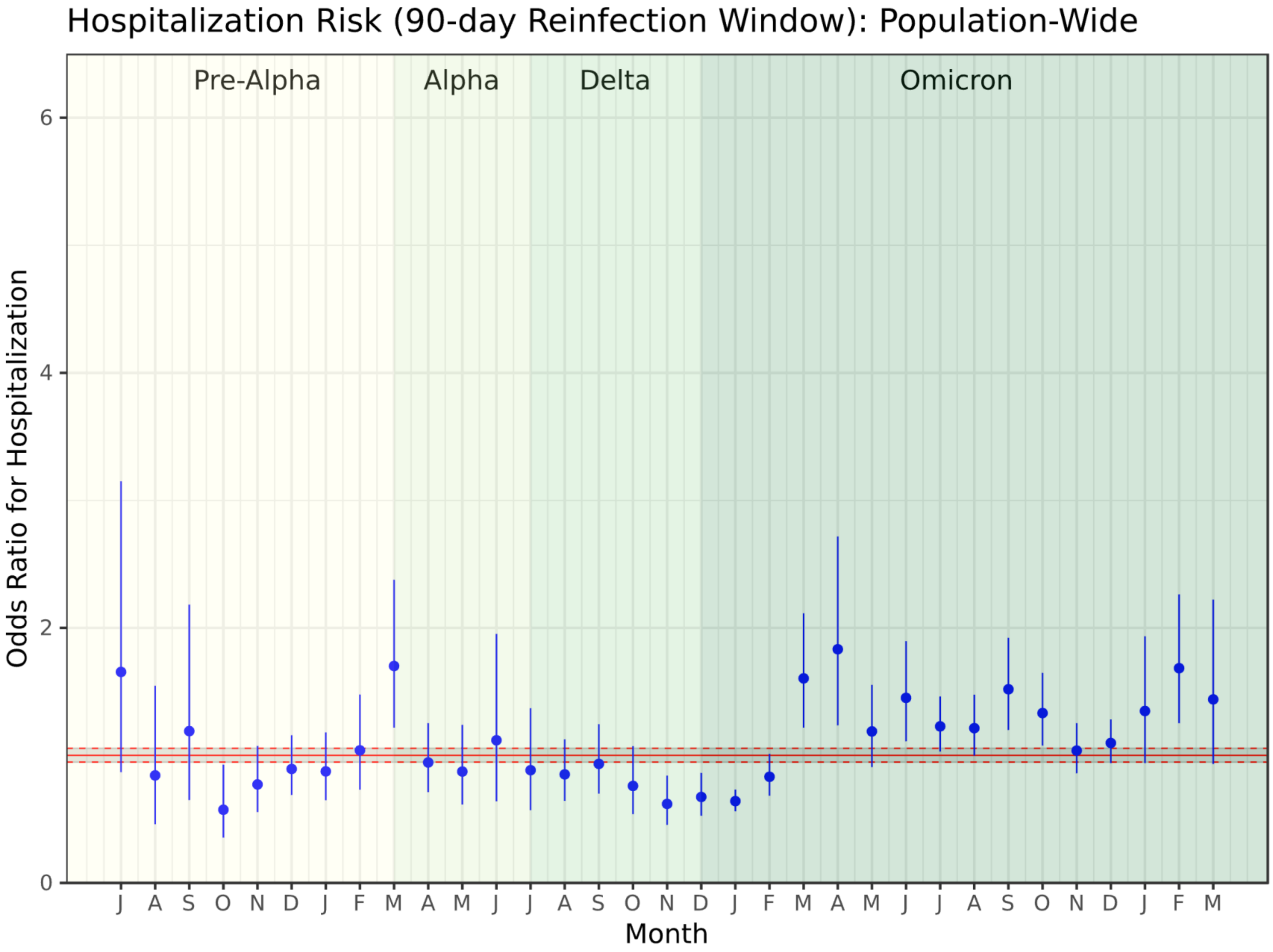

VII. Risk of death: No lookback period

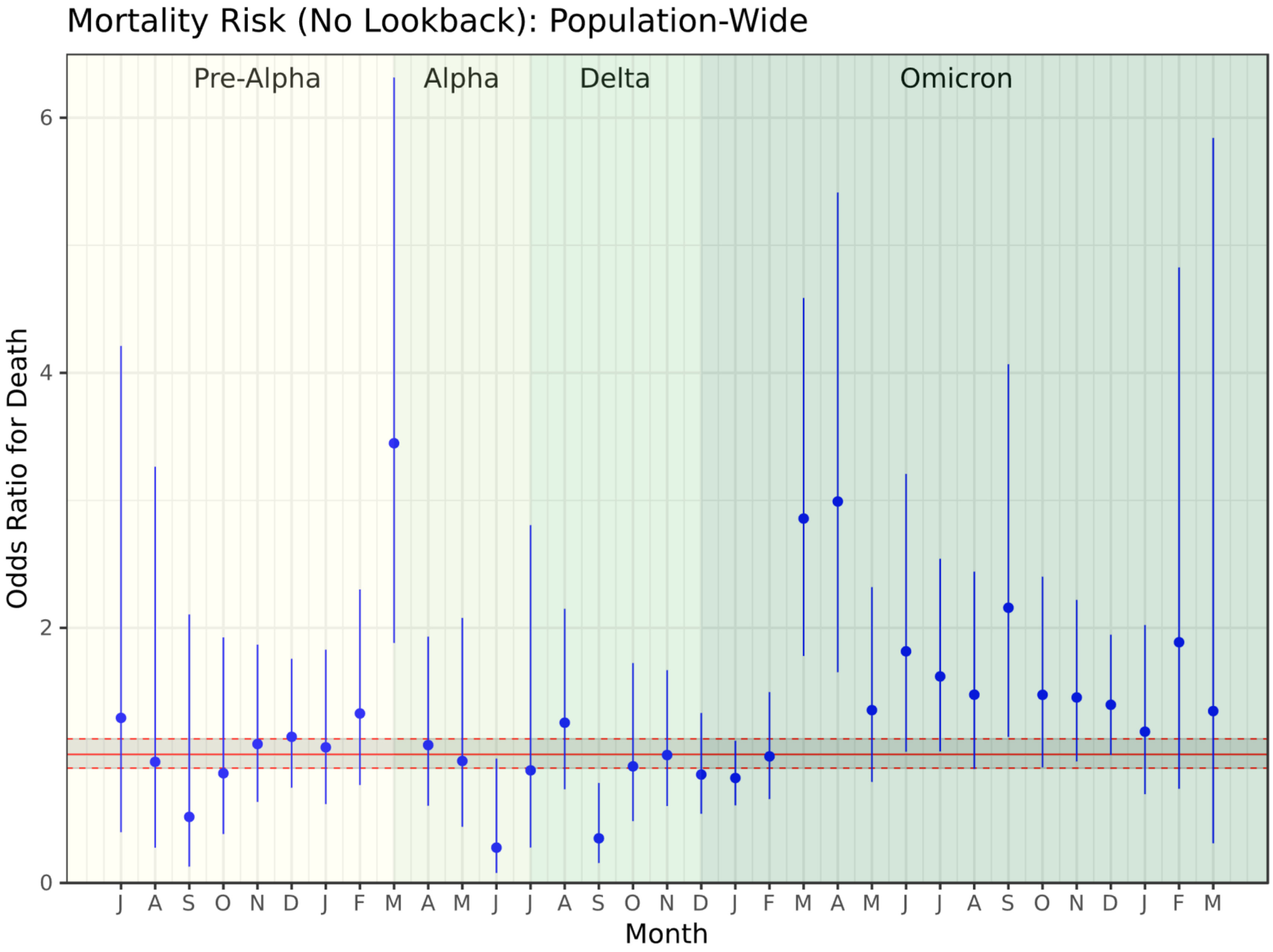

VIII. Risk of death: 90-day reinfection window

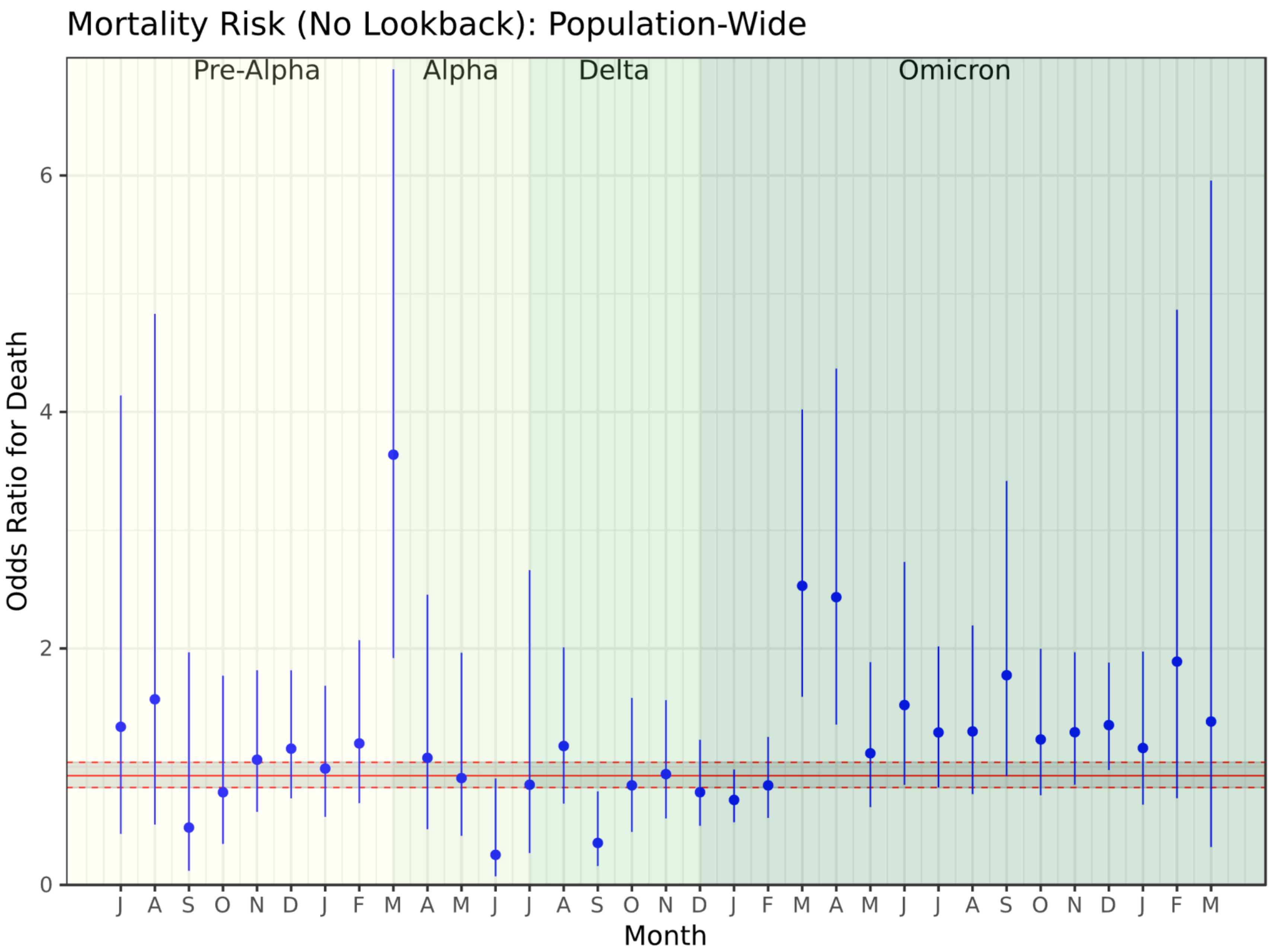

IX. Subgroup analyses of overall severity by race/ethnicity, gender, and age

*Pandemic-wide and minimum/maximum odds ratios (ORs) for more severe disease*

| | Pandemic-Wide OR (95% CI) | Minimum Monthly OR (95% CI) | Maximum Monthly OR (95% CI) |
|---|---|---|---|
| **Full Cohort** | 1.06 (1.03 – 1.10) | October 2020: 0.56 (0.37 – 0.84) | February 2023: 1.64 (1.33 – 2.00) |
| **Black / African American** | 1.08 (1.01 – 1.15) | November 2020: 0.36 (0.15 – 0.86) | March 2021: 1.64 (0.69 – 3.91) |
| **Hispanic** | 0.99 (0.92 – 1.08) | January 2021: 0.51 (0.18 – 1.47) | July 2020: 1.64 (0.49 – 5.51) |
| **White** | 1.02 (0.98 – 1.07) | August 2020: 0.12 (0.04 – 0.39) | February 2023: 2.01 (1.49 – 2.70) |
| **Female** | 1.00 (0.96 – 1.04) | November 2020: 0.61 (0.42 – 0.87) | September 2020: 1.79 (0.89 – 3.60) |
| **Male** | 1.11 (1.06 – 1.16) | October 2020: 0.29 (0.14 – 0.64) | February 2023: 1.79 (1.24 – 2.58) |
| **Age <18** | 1.34 (1.22 – 1.49) | February 2021: 0.49 (0.07 – 3.56) | June 2021: 2.56 (0.58 – 11.22) |
| **Age 18 – 44** | 0.99 (0.95 – 1.03) | November 2020: 0.44 (0.21 – 0.90) | September 2020: 2.34 (0.85 – 6.45) |
| **Age 45 – 59** | 0.92 (0.87 – 0.97) | August 2020: 0.11 (0.03 – 0.39) | February 2023: 1.96 (1.21 – 3.16) |
| **Age 60 – 69** | 0.92 (0.86 – 0.99) | June 2021: 0.28 (0.11 – 0.71) | March 2022: 1.62 (0.86 – 3.04) |
| **Age ≤70** | 0.89 (0.85 – 0.94) | July 2020: 0.24 (0.09 – 0.65) | April 2022: 2.00 (1.32 – 3.03) |

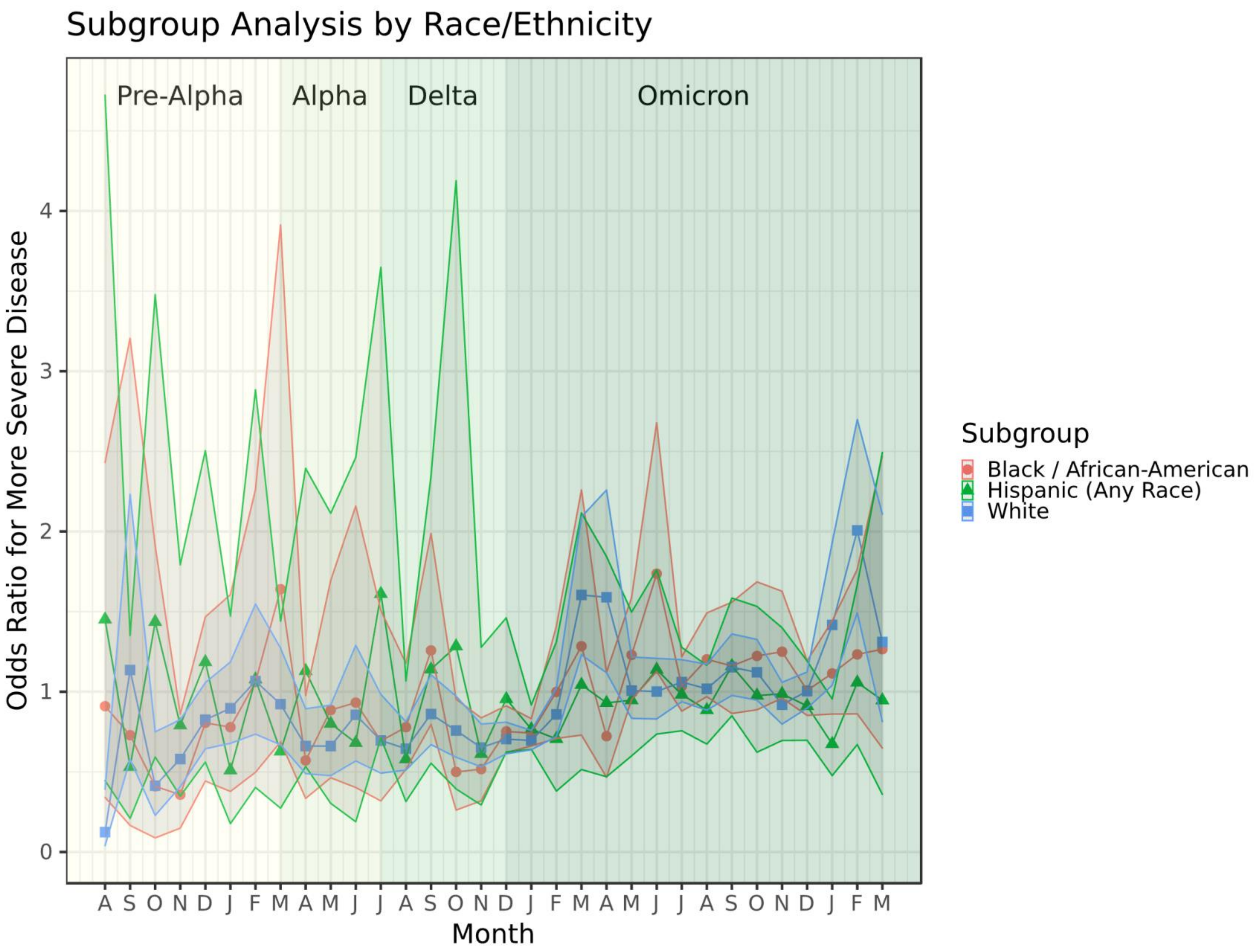
Subgroup Analysis by Race/Ethnicity
Pre-Alpha
Alpha
Delta
Omicron
Odds Ratio for More Severe Disease
0
1
2
3
4
A S O N D J F M A M J J A S O N D J F M A M J J A S O N D J F M
Month
Subgroup
Black / African-American
Hispanic (Any Race)
White

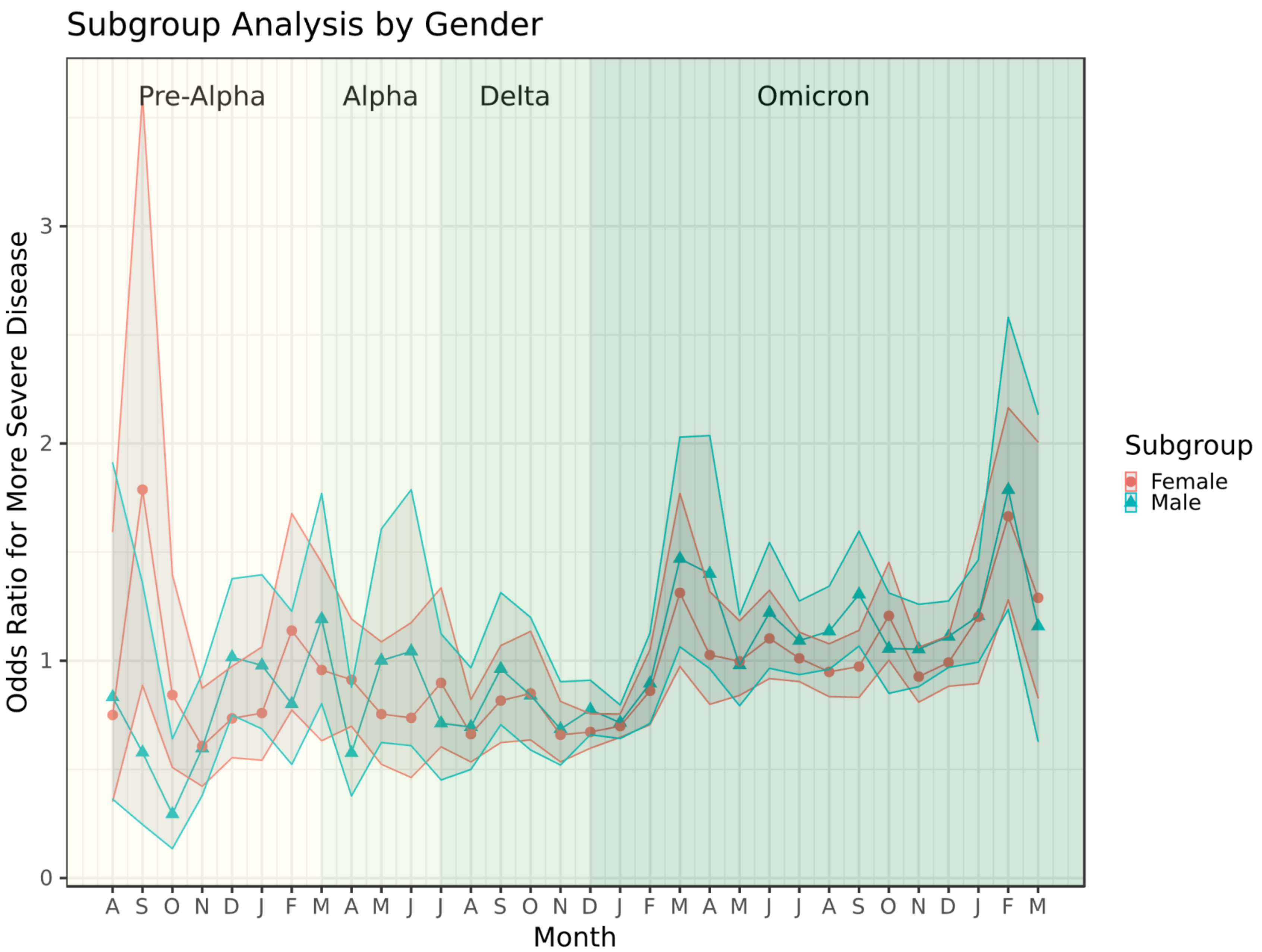
Subgroup Analysis by Gender
Pre-Alpha
Alpha
Delta
Omicron
Odds Ratio for More Severe Disease
0
1
2
3
A S O N D J F M A M J J A S O N D J F M A M J J A S O N D J F M
Month
Subgroup
Female
Male

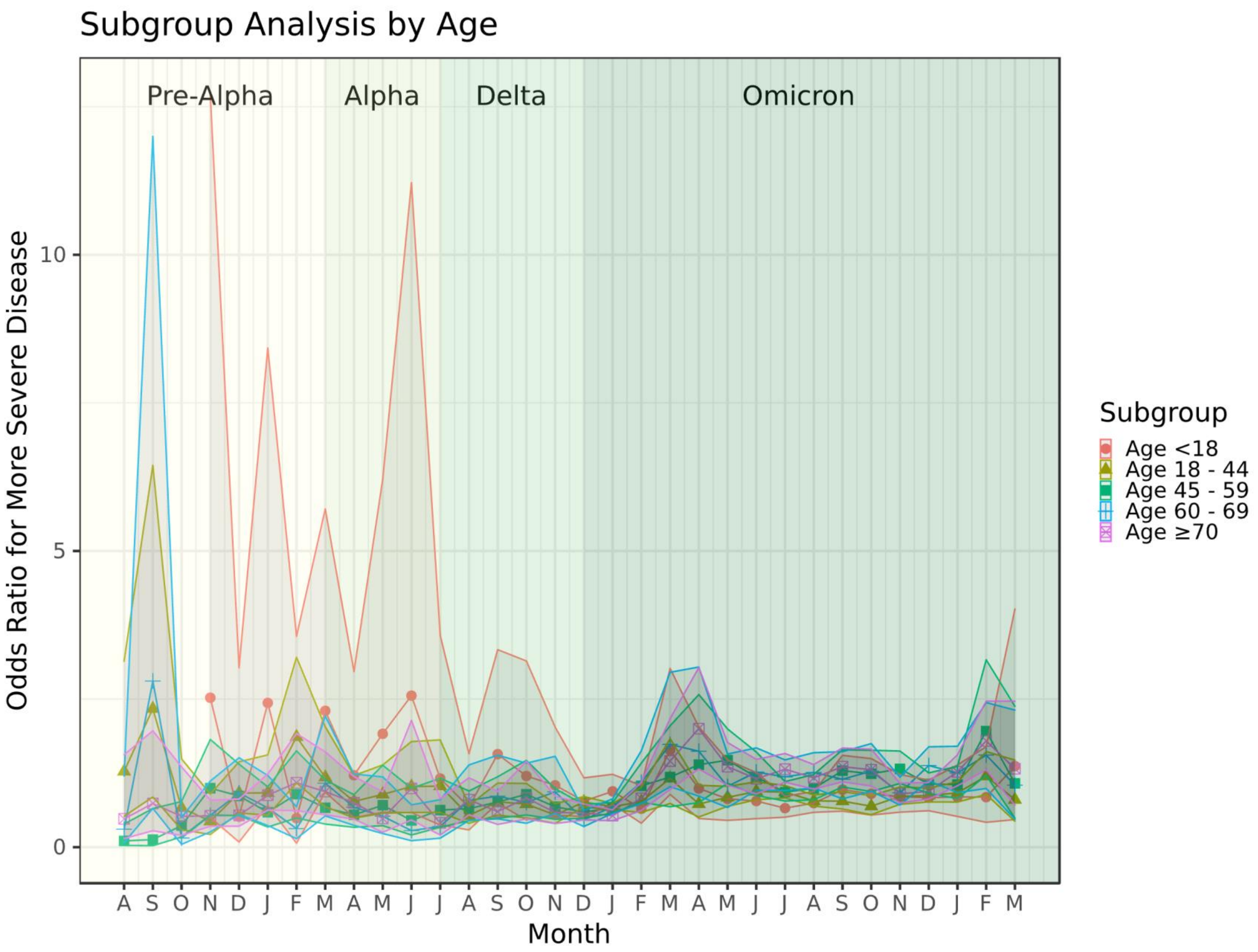
Subgroup Analysis by Age
Pre-Alpha
Alpha
Delta
Omicron
Odds Ratio for More Severe Disease
10
5
0
A S O N D J F M A M J J A S O N D J F M A M J J A S O N D J F M
Month
Subgroup
Age <18
Age 18 - 44
Age 45 - 59
Age 60 - 69
Age ≥70

X. <u>Overall severity: No visit within lookback period required for included patients</u>

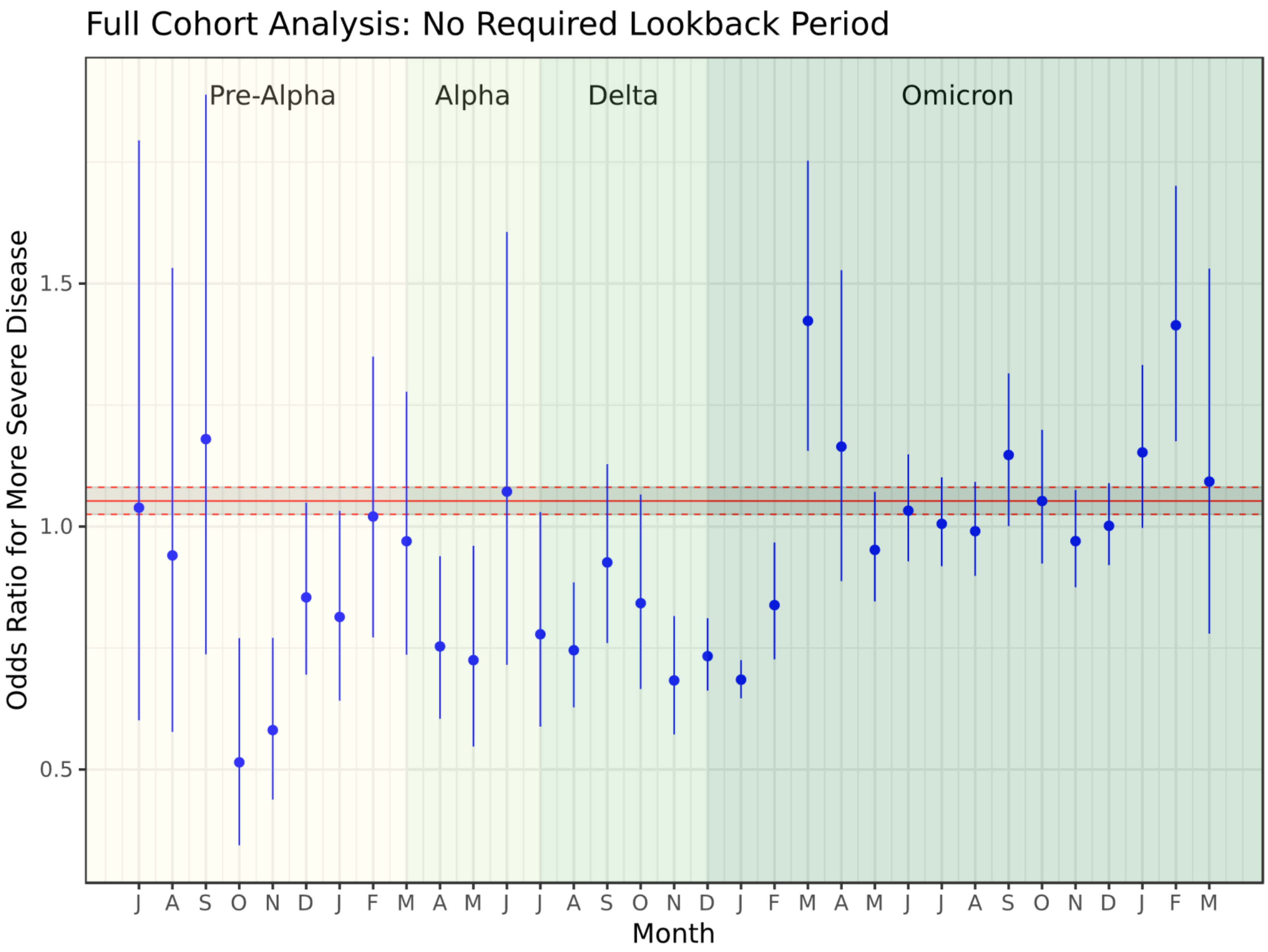

XI. Overall severity: Minimum of 90 days required between first infection and second infection

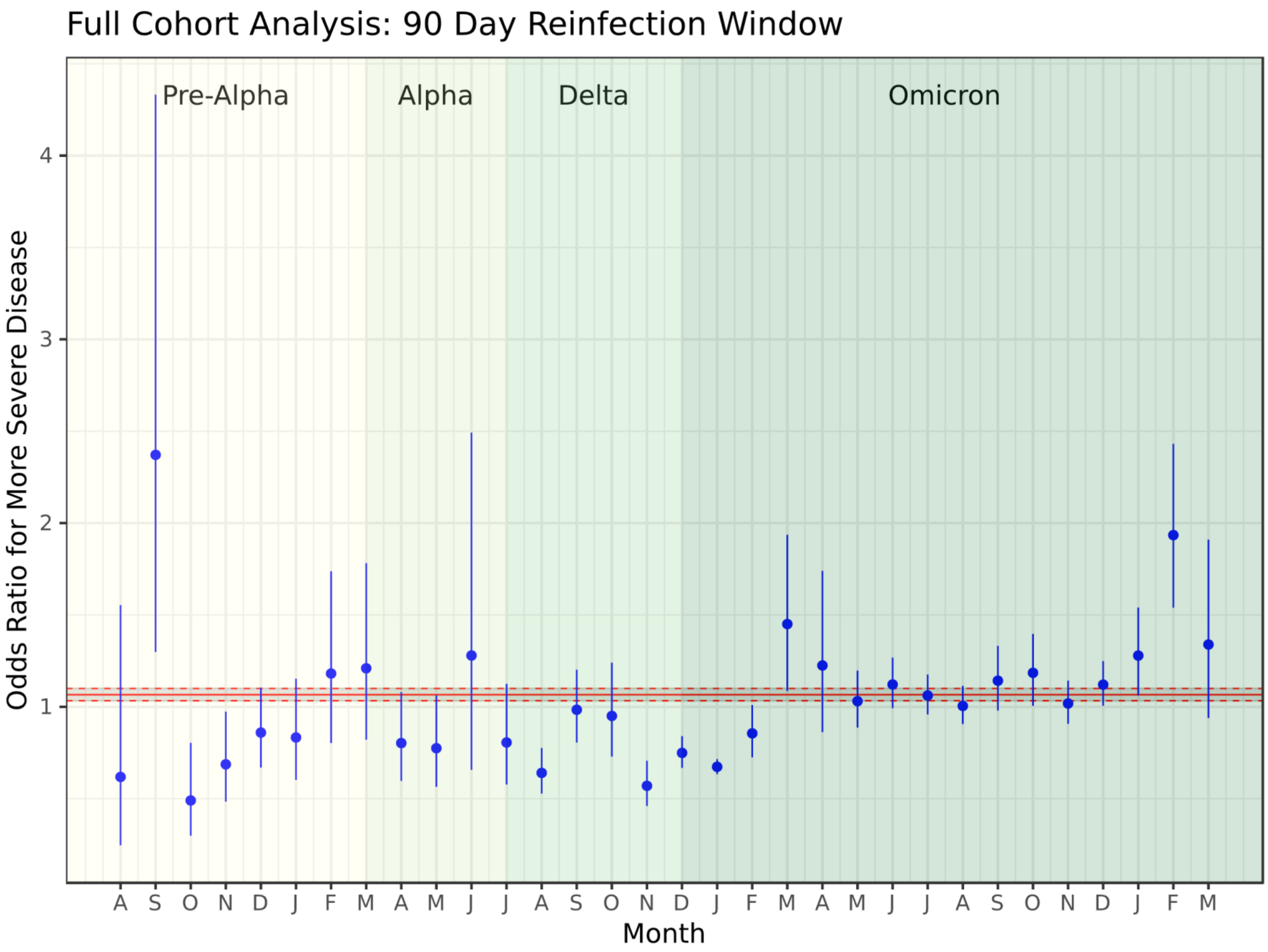

**Individual Acknowledgements for Core Contributors:** We gratefully acknowledge the following core contributors to N3C: Adam B. Wilcox, Adam M. Lee, Alexis Graves, Alfred (Jerrod) Anzalone, Amin Manna, Amit Saha, Amy Olex, Andrea Zhou, Andrew E. Williams, Andrew Southerland, Andrew T. Girvin, Anita Walden, Anjali A. Sharathkumar, Benjamin Amor, Benjamin Bates, Brian Hendricks, Brijesh Patel, Caleb Alexander, Carolyn Bramante, Cavin Ward-Caviness, Charisse Madlock-Brown, Christine Suver, Christopher Chute, Christopher Dillon, Chunlei Wu, Clare Schmitt, Cliff Takemoto, Dan Housman, Davera Gabriel, David A. Eichmann, Diego Mazzotti, Don Brown, Eilis Boudreau, Elaine Hill, Elizabeth Zampino, Emily Carlson Marti, Emily R. Pfaff, Evan French, Farrukh M Koraishy, Federico Mariona, Fred Prior, George Sokos, Greg Martin, Harold Lehmann, Heidi Spratt, Hemalkumar Mehta, Hongfang Liu, Hythem Sidky, J.W. Awori Hayanga, Jami Pincavitch, Jaylyn Clark, Jeremy Richard Harper, Jessica Islam, Jin Ge, Joel Gagnier, Joel H. Saltz, Joel Saltz, Johanna Loomba, John Buse, Jomol Mathew, Joni L. Rutter, Julie A. McMurry, Justin Guinney, Justin Starren, Karen Crowley, Katie Rebecca Bradwell, Kellie M. Walters, Ken Wilkins, Kenneth R. Gersing, Kenrick Dwain Cato, Kimberly Murray, Kristin Kostka, Lavance Northington, Lee Allan Pyles, Leonie Misquitta, Lesley Cottrell, Lili Portilla, Mariam Deacy, Mark M. Bissell, Marshall Clark, Mary Emmett, Mary Morrison Saltz, Matvey B. Palchuk, Melissa A. Haendel, Meredith Adams, Meredith Temple-O'Connor, Michael G. Kurilla, Michele Morris, Nabeel Qureshi, Nasia Safdar, Nicole Garbarini, Noha Sharafeldin, Ofer Sadan, Patricia A. Francis, Penny Wung Burgoon, Peter Robinson, Philip R.O. Payne, Rafael Fuentes, Randeep Jawa, Rebecca Erwin-Cohen, Rena Patel, Richard A. Moffitt, Richard L. Zhu, Rishi Kamaleswaran, Robert Hurley, Robert T. Miller, Saiju Pyarajan, Sam G. Michael, Samuel Bozzette, Sandeep Mallipattu, Satyanarayana Vedula, Scott Chapman, Shawn T. O'Neil, Soko Setoguchi, Stephanie S. Hong, Steve Johnson, Tellen D. Bennett, Tiffany Callahan, Umit Topaloglu, Usman Sheikh, Valery Gordon, Vignesh Subbian, Warren A. Kibbe, Wenndy Hernandez, Will Beasley, Will Cooper, William Hillegass, Xiaohan Tanner Zhang. Details of contributions available at covid.cd2h.org/core-contributors

**Data Partners with Released Data: The following institutions whose data is released or pending:**
Available: Advocate Health Care Network — UL1TR002389: The Institute for Translational Medicine (ITM) • Boston University Medical Campus — UL1TR001430: Boston University Clinical and Translational Science Institute • Brown University — U54GM115677: Advance Clinical Translational Research (Advance-CTR) • Carilion Clinic — UL1TR003015: iTHRIV Integrated Translational health Research Institute of Virginia • Charleston Area Medical Center — U54GM104942: West Virginia Clinical and Translational Science Institute (WVCTSI) • Children's Hospital Colorado — UL1TR002535: Colorado Clinical and Translational Sciences Institute • Columbia University Irving Medical Center — UL1TR001873: Irving Institute for Clinical and Translational Research • Duke University — UL1TR002553: Duke Clinical and Translational Science Institute • George Washington Children's Research Institute — UL1TR001876: Clinical and Translational Science Institute at Children's National (CTSA-CN) • George Washington University — UL1TR001876: Clinical and Translational Science Institute at Children's National (CTSA-CN) • Indiana University School of Medicine — UL1TR002529: Indiana Clinical and Translational Science Institute • Johns Hopkins University — UL1TR003098: Johns Hopkins Institute for Clinical and Translational Research • Loyola Medicine — Loyola University Medical Center • Loyola University Medical Center — UL1TR002389: The Institute for Translational Medicine (ITM) • Maine Medical Center — U54GM115516: Northern New England Clinical & Translational Research (NNE-CTR) Network • Massachusetts General Brigham — UL1TR002541: Harvard Catalyst • Mayo Clinic Rochester — UL1TR002377: Mayo Clinic Center for Clinical and Translational Science (CCaTS) • Medical University of South Carolina — UL1TR001450: South Carolina Clinical & Translational Research Institute (SCTR) • Montefiore Medical Center — UL1TR002556: Institute for Clinical and Translational Research at Einstein and Montefiore • Nemours — U54GM104941: Delaware CTR ACCEL Program • NorthShore University HealthSystem — UL1TR002389: The Institute for Translational Medicine (ITM) • Northwestern University at Chicago — UL1TR001422: Northwestern University Clinical and Translational Science Institute (NUCATS) • OCHIN — INV-018455: Bill and Melinda Gates Foundation grant to Sage Bionetworks • Oregon Health & Science University — UL1TR002369: Oregon Clinical and Translational Research Institute • Penn State Health Milton S. Hershey Medical Center — UL1TR002014: Penn State Clinical and Translational Science Institute • Rush University Medical Center —

UL1TR002389: The Institute for Translational Medicine (ITM) • Rutgers, The State University of New Jersey — UL1TR003017: New Jersey Alliance for Clinical and Translational Science • Stony Brook University — U24TR002306 • The Ohio State University — UL1TR002733: Center for Clinical and Translational Science • The State University of New York at Buffalo — UL1TR001412: Clinical and Translational Science Institute • The University of Chicago — UL1TR002389: The Institute for Translational Medicine (ITM) • The University of Iowa — UL1TR002537: Institute for Clinical and Translational Science • The University of Miami Leonard M. Miller School of Medicine — UL1TR002736: University of Miami Clinical and Translational Science Institute • The University of Michigan at Ann Arbor — UL1TR002240: Michigan Institute for Clinical and Health Research • The University of Texas Health Science Center at Houston — UL1TR003167: Center for Clinical and Translational Sciences (CCTS) • The University of Texas Medical Branch at Galveston — UL1TR001439: The Institute for Translational Sciences • The University of Utah — UL1TR002538: Uhealth Center for Clinical and Translational Science • Tufts Medical Center — UL1TR002544: Tufts Clinical and Translational Science Institute • Tulane University — UL1TR003096: Center for Clinical and Translational Science • University Medical Center New Orleans — U54GM104940: Louisiana Clinical and Translational Science (LA CaTS) Center • University of Alabama at Birmingham — UL1TR003096: Center for Clinical and Translational Science • University of Arkansas for Medical Sciences — UL1TR003107: UAMS Translational Research Institute • University of Cincinnati — UL1TR001425: Center for Clinical and Translational Science and Training • University of Colorado Denver, Anschutz Medical Campus — UL1TR002535: Colorado Clinical and Translational Sciences Institute • University of Illinois at Chicago — UL1TR002003: UIC Center for Clinical and Translational Science • University of Kansas Medical Center — UL1TR002366: Frontiers: University of Kansas Clinical and Translational Science Institute • University of Kentucky — UL1TR001998: UK Center for Clinical and Translational Science • University of Massachusetts Medical School Worcester — UL1TR001453: The UMass Center for Clinical and Translational Science (UMCCTS) • University of Minnesota — UL1TR002494: Clinical and Translational Science Institute • University of Mississippi Medical Center — U54GM115428: Mississippi Center for Clinical and Translational Research (CCTR) • University of Nebraska Medical Center — U54GM115458: Great Plains IDeA-Clinical & Translational Research • University of North Carolina at Chapel Hill — UL1TR002489: North Carolina Translational and Clinical Science Institute • University of Oklahoma Health Sciences Center — U54GM104938: Oklahoma Clinical and Translational Science Institute (OCTSI) • University of Rochester — UL1TR002001: UR Clinical & Translational Science Institute • University of Southern California — UL1TR001855: The Southern California Clinical and Translational Science Institute (SC CTSI) • University of Vermont — U54GM115516: Northern New England Clinical & Translational Research (NNE-CTR) Network • University of Virginia — UL1TR003015: iTHRIV Integrated Translational health Research Institute of Virginia • University of Washington — UL1TR002319: Institute of Translational Health Sciences • University of Wisconsin-Madison — UL1TR002373: UW Institute for Clinical and Translational Research • Vanderbilt University Medical Center — UL1TR002243: Vanderbilt Institute for Clinical and Translational Research • Virginia Commonwealth University — UL1TR002649: C. Kenneth and Dianne Wright Center for Clinical and Translational Research • Wake Forest University Health Sciences — UL1TR001420: Wake Forest Clinical and Translational Science Institute • Washington University in St. Louis — UL1TR002345: Institute of Clinical and Translational Sciences • Weill Medical College of Cornell University — UL1TR002384: Weill Cornell Medicine Clinical and Translational Science Center • West Virginia University — U54GM104942: West Virginia Clinical and Translational Science Institute (WVCTSI)

Submitted: Icahn School of Medicine at Mount Sinai — UL1TR001433: ConduITS Institute for Translational Sciences • The University of Texas Health Science Center at Tyler — UL1TR003167: Center for Clinical and Translational Sciences (CCTS) • University of California, Davis — UL1TR001860: UCDavis Health Clinical and Translational Science Center • University of California, Irvine — UL1TR001414: The UC Irvine Institute for Clinical and Translational Science (ICTS) • University of California, Los Angeles — UL1TR001881: UCLA Clinical Translational Science Institute • University of California, San Diego — UL1TR001442: Altman Clinical and Translational Research Institute • University of California, San Francisco — UL1TR001872: UCSF Clinical and Translational Science Institute

Pending: Arkansas Children's Hospital — UL1TR003107: UAMS Translational Research Institute • Baylor College of Medicine — None (Voluntary) • Children's Hospital of Philadelphia — UL1TR001878: Institute for Translational Medicine and Therapeutics • Cincinnati Children's Hospital Medical Center — UL1TR001425: Center for Clinical and Translational Science and Training • Emory University — UL1TR002378: Georgia Clinical and Translational Science Alliance • HonorHealth — None (Voluntary) • Loyola University Chicago — UL1TR002389: The Institute for Translational Medicine (ITM) • Medical College of Wisconsin — UL1TR001436: Clinical and Translational Science Institute of Southeast Wisconsin • MedStar Health Research Institute — UL1TR001409: The Georgetown-Howard Universities Center for Clinical and Translational Science (GHUCCTS) • MetroHealth — None (Voluntary) • Montana State University — U54GM115371: American Indian/Alaska Native CTR • NYU Langone Medical Center — UL1TR001445: Langone Health's Clinical and Translational Science Institute • Ochsner Medical Center — U54GM104940: Louisiana Clinical and Translational Science (LA CaTS) Center • Regenstrief Institute — UL1TR002529: Indiana Clinical and Translational Science Institute • Sanford Research — None (Voluntary) • Stanford University — UL1TR003142: Spectrum: The Stanford Center for Clinical and Translational Research and Education • The Rockefeller University — UL1TR001866: Center for Clinical and Translational Science • The Scripps Research Institute — UL1TR002550: Scripps Research Translational Institute • University of Florida — UL1TR001427: UF Clinical and Translational Science Institute • University of New Mexico Health Sciences Center — UL1TR001449: University of New Mexico Clinical and Translational Science Center • University of Texas Health Science Center at San Antonio — UL1TR002645: Institute for Integration of Medicine and Science • Yale New Haven Hospital — UL1TR001863: Yale Center for Clinical Investigation
